# Guillain-Barré syndrome following Zika virus infection is associated with a diverse spectrum of peripheral nerve reactive antibodies

**DOI:** 10.1101/2021.10.28.21265167

**Authors:** Alexander J Davies, Cinta Lleixà, Ana M. Siles, Dawn Gourlay, Georgina Berridge, Wanwisa Dejnirattisai, Carolina Ramírez-Santana, Juan-Manuel Anaya, Andrew K. Falconar, Claudia M. Romero-Vivas, Lyda Osorio, Beatriz Parra, Gavin R. Screaton, Juthathip Mongkolsapaya, Roman Fischer, Carlos A. Pardo, Susan K. Halstead, Hugh J. Willison, Luis Querol, Simon Rinaldi

## Abstract

**Introduction:** Recent outbreaks of Zika virus (ZIKV) in South and Central America have highlighted significant neurological side effects. Concurrence with the inflammatory neuropathy Guillain-Barré syndrome (GBS) is observed in 1:4000 ZIKV cases. Whether the neurological symptoms of ZIKV infection are a consequence of autoimmunity or direct neurotoxicity is unclear.

**Methods:** We employed rat dorsal root ganglion (DRG) neurons, Schwann cells (SCs), and human stem cell-derived sensory neurons myelinated with rat SCs as cellular models to screen for IgG and IgM autoantibodies reactive to peripheral nerve in sera of ZIKV patients with and without GBS. In this study, 52 ZIKV-GBS patients were compared with 134 ZIKV-infected patients, and 91 non-ZIKV controls. Positive sera were taken forward for target identification by immunoprecipitation and mass spectrometry, and candidate antigens validated by ELISA and cell-based assays. Autoantibody reactions against glycolipid antigens were also screened on an array.

**Results:** Overall, IgG antibody reactivity to rat SCs (6.5%) and myelinated co-cultures (9.6%) were significantly higher, albeit infrequently, in the ZIKV-GBS group compared to all controls. IgM antibody immunoreactivity to DRGs (32.3%) and SCs (19.4%) was more frequently observed in the ZIKV-GBS group compared to other controls, while IgM reactivity to co-cultures was as common in ZIKV and non-ZIKV sera. Strong axonal-binding ZIKV-GBS serum IgG antibodies from one patient were confirmed to react with neurofascin-155 and 186. Serum from a ZIKV non-GBS patient displayed strong myelin-binding and anti-lipid antigen reaction characteristics. There was no significant association of ZIKV-GBS with any anti-glycolipid antibodies.

**Conclusion:** Autoantibodies in ZIKV associated GBS patients’ sera target heterogeneous peripheral nerve antigens suggesting heterogeneity of the humoral immune response despite a common prodromal infection.

## INTRODUCTION

Zika virus (ZIKV) is a member of the Flavivirus family that is transmitted by *Aedes* spp. mosquito vector species and in humans typically results in asymptomatic or mildly symptomatic infections. An outbreak of ZIKV infections in French Polynesia in 2013 was followed by a spike in the diagnoses of the disabling, acute inflammatory neuropathy Guillain-Barré syndrome (GBS). Early case control studies established one excess GBS case for every 4000 people infected with ZIKV.[1] By 2015, the ZIKV pandemic reached South America, where in Colombia the GBS incidence rates were increased by 210-350% compared to the pre-ZIKV period.[2, 3] The overall risk estimates range from 1 to 8 GBS cases per 10,000 ZIKV infections.[4] Recent studies estimate that the total number of ZIKV infections during the 2015-16 outbreak was much higher than originally reported, suggesting previous incidence rates of GBS per infection may have been substantially underestimated.[5]

The onset of GBS post-ZIKV infection (ZIKV-GBS) occurred within a median of 6 days after a transient febrile illness, though the peak incidence of GBS cases was observed 3 weeks after the peak of ZIKV diagnoses.[1, 2] Whether this represents an infectious, para-infectious or post-infectious aetiology has been debated. Electrophysiological studies performed on 37 ZIKV-GBS patients identified in French Polynesia showed characteristics consistent with the acute motor axonal form of GBS (AMAN).[1] However, follow-up studies at 4 months were more suggestive of distal demyelination.[6] In a Colombian series, 70% of ZIKV-GBS cases were classified as acute inflammatory demyelinating polyradiculoneuropathy (AIDP), suggestive of immune-mediated injury.[7] ZIKV has also been linked to a number of central and peripheral neurological complications, all of which may have an autoimmune mechanism [8].

A potential role for ZIKV-induced autoimmunity is supported by similarities between the ZIKV E protein and human complement component C1q *in silico*, and the presence of C1q antibodies in the sera of animals infected with the virus.[9] Lynch and colleagues observed higher-titre antibody responses to ZIKV in patients who developed GBS, compared to those with uncomplicated infections.[10] More recently, a higher incidence of anti-ganglioside antibodies was identified in a Brazilian cohort of ZIKV-GBS patients compared to uncomplicated ZIKV controls [11] suggesting an association between nerve reactive autoantibodies and the peripheral neurological complications of ZIKV.

The peripheral neurology associated with ZIKV infection may also occur via direct viral toxicity.[12] ZIKV directly infected peripheral neurons and induced cell death in both mouse and cell culture models.[13], as well as infecting sensory neurons and Schwann cells (SCs) in dorsal root ganglia (DRG) explants from interferon receptor-1 knockout mice leading to myelin fragmentation and later axonal degeneration.[14]

In this study, we sought to address the potential for humoral autoimmunity in ZIKV-GBS by screening sera from a large cohort of ZIKV-GBS patients and controls for nerve reactive immunoglobulins using a range of animal and humanised nerve cell cultures and antigen detection methods.

## MATERIALS & METHODS

Full details of this section are available in Supplementary Material and Data files.

### Patient cohorts and sample acquisition

Serum samples were collected from patients with ZIKV infection both with and without neurological complications, as well as from other infectious, healthy and convalescent controls from the Cucuta, Cali and Barranquilla regions of Colombia, the UK (South Central - Oxford A approval number 14/SC/0280) and Spain (The Sant Pau Biomedical Research Institute approval number IIBSP-AUT-2016-69) (**Supplementary Figure 1** and **Supplementary Table 1**). ZIKV-GBS patients were classified according to previously published electrodiagnostic criteria, [7] with full characterisations previously reported for Cucuta [15] and Cali cohorts. [3] Serum samples were aliquoted and stored at −80°C. This study was carried out and reported in accordance with the STROBE guidelines.[16]

### Rat DRG neuron and SC culture and immunocytochemistry

Rat DRG neurons and SCs were extracted in accordance with Schedule 1 of the UK Home Office Animals (Scientific Procedures) Act 1986, and Animal Ethics’ Committee of Hospital de la Santa Creu i Sant Pau. Cells were cultured, and immunocytochemistry was performed as previously described.[17] Fluorescence signal intensities were scored on a 0–3 scale by two independent researchers.

### Myelinating co-cultures and immunofluorescence labelling

Myelinating co-cultures were prepared using human induced pluripotent stem cell (hiPSC)-derived sensory neurons and neonatal rat SCs as previously described.[18, 19] Sera from ZIKV-exposed subjects was diluted to 1:100 and added to live co-cultures for 1h at 37°C. Fixed cultures were probed with either anti-human IgG, anti-human IgM or anti-human complement C3c antibodies. Confocal images were assessed for IgG or IgM reactivity by a blinded observer.

### Assessment of serum-induced demyelination

Myelinating co-cultures were incubated for 1h in ‘complete’ neurobasal media supplemented with fluoromyelin red. Serum-free myelination medium was then supplemented with or without human anti-ZIKV patients’ sera at a 1:100 dilution together with 20% normal human serum (NHS) as a source of complement and added to the co-cultures.

### Glycoarray

Serum samples were also screened on a glycolipid microarray as previously described.[20] For this study, sera were screened against a panel of 16 single glycolipids (GM1, GM2, phosphatidylserine, GM4, GA1, GD1a, GD1b, GT1a, GT1b, GQ1b, GD3, SGPG, LM1, cholesterol, GalC and sulphatide) and 120 heteromeric 1:1 (v:v) complexes, printed in duplicate.

### Immunoprecipitation and mass spectrometry

Human sera showing moderate or strong reactivity against rat DRG neurons or myelinating co-cultures were used for immunoprecipitation (IP) experiments using the same target cell type, as previously reported.[17] For the mass spectrometry analyses, the IP eluates from myelinating co-cultures were prepared by chloroform:methanol precipitation and in-solution trypsin digestion. The resultant peptides were then subjected to high resolution C18 reverse-phase column chromatography and analysed by data-dependant MS/MS on a ThermoFisher Fusion Lumos mass spectrometer. Raw data are available via ProteomeXchange with identifier PXD028476.

### Protein electrophoresis and Western blot

The lysates from myelinating co-cultures, hiPSC-derived sensory neurons monocultures, and primary rat SCs were subjected to electrophoresis in acrylamide gels using MOPS running buffer under denaturing, non-reducing conditions and then transferred to a nitrocellulose membrane. These blots were probed with using patients’ sera at a 1:2500 dilution followed by HRP-conjugated Fc-specific anti-human-IgG secondary antibody. The bands detected by western blotting were aligned in a parallel gel stained with Pierce Imperial Protein Stain and excised for mass spectrometry.

### ELISA

Recombinant human nidogen-1, laminins 111, 121, 211, 221, 411, 421, 511 and 521, prosaposin and vinculin were coated at 1 μg/ml (100 μl per well) in Maxisorp ELISA plates overnight at 4°C (See **Supplementary Table 2** for details). The ganglioside GM3 ELISA was performed as previously described [21]. For the detection of anti-ZIKV antibodies, Maxisorp plates were coated with the anti-flavivirus mouse monoclonal antibody 4G2 and performed as previously described.[22] Bound human IgG antibodies were detected through sequential steps using Fc-specific HRP-conjugated secondary antibodies and substrate.

### Transfected-cell based assays

HEK293 cells were transfected overnight using JetPEI with mammalian expression vectors encoding human AHNAK2, ANXA2, CD44S, CNTN1, CASPR1, GFRA1, ITGA6, ITGA7, MAG, NEP, NFASC, NKCC1, PRX, TGBR3 (**Supplementary Table 3**), or using Lipofectamine 2000 for human ALCAM, AXL, DPYSL2, GAS6, L1CAM, NCAM1 and NrCAM (**Supplementary Table 4**). Sera were diluted 1:100 and IgG binding was assessed using fluorescent-conjugated anti-human IgG secondary antibodies.

### Statistical analysis

The results were analysed in Prism v9.1.0 (GraphPad). Statistical comparisons of the proportions of seropositive patients among ZIKV groups were performed using contingency analyses with the application of a two-tailed Fisher’s exact test for individual group-group comparisons, and Chi square testing for comparisons between all groups.

## RESULTS

### Patient cohorts

Serum samples were obtained from a total of 218 Colombian subjects: 52 patients with GBS and confirmed ZIKV infection (ZIKV-GBS), 17 patients with ZIKV infection and other inflammatory neurological diseases (ZIKV-OND), 117 ZIKV-infected patients without neurological complications (ZIKV-CON), and a control group (CON) formed by 23 patients with Dengue virus (DENV) infections (DENV-CON) from Cali and 9 healthy controls from Cucuta. An additional 38 serum samples (5 ALS and 5 Dysferlin patients, and 28 healthy controls) from Spanish subjects and 21 healthy controls from the UK were also used (**Supplementary Figure 1).**

Overall, the patients with ZIKV-GBS were slightly older and less often female (**Supplementary Table 1**). ZIKV capture ELISA showed the presence of anti-ZIKV IgG in all ZIKV-GBS, ZIKV-OND and ZIKV-CON patients’ sera tested, as well as the DENV patients’ sera, probably due to the likely cross-reactivity of DENV and ZIKV patients’ antibodies [22] (**Supplementary Figure 2**).

### Microarray screening for glycolipid complex antibodies

ZIKV-associated patients’ sera were screened against a panel of glycolipids using a combinatorial glycoarray for the presence of IgG (**Supplementary Figure 3A**) and IgM (**Supplementary Figure 3B**) anti-glycolipid antibodies.

Overall, the glycolipid array revealed only weak binding intensities against the panel of 136 unique glycolipid targets, including known gangliosides implicated in GBS, none of which were found to be statistically associated with post-ZIKV GBS. Two ZIKV-GBS samples were found to contain IgG antibodies against various gangliosides at elevated levels, as typically seen in GBS and its clinical variants. This included a single ZIKV-GBS sample with anti-disialosyl IgG (GQ1b, GT1a and GD3) normally associated with the GBS subtype Miller-Fisher syndrome; another with anti-GM1 IgG, typically present in the clinical subtype AMAN. It is possible that these cases may represent a temporal co-infection of Campylobacter and Flavivirus, resulting in a classic IgG antibody signature for these patients.

### Screening against rat DRG neurons and Schwann cells

Moderate or strongly rat DRG reactive IgM autoantibodies were observed in proportionally more sera of ZIKV-GBS patients compared to the ZIKV-OND, ZIKV-CON patients and other controls (CON) groups (**Table 1**). On the other hand, moderate or strong IgG autoantibody reactivity against DRG neurons were not statistically different between these groups. With the SCs, a greater proportion of moderate or strong IgG and IgM reactivity was observed in ZIKV-GBS patients’ sera compared to other uncomplicated ZIKV patients and control groups (**Table 1**).

**Table 1.** Statistical analysis of serum immunoreactivity to DRG neurons and SC cultures.

|  |  | ZIKV - GBS<br>(n = 31) |  | ZIKV-OND<br>(n = 12) |  | ZIKV-CON<br>(n = 77) |  | CON<br>(n = 43) |  | Chi-square<br>p value |  |
| --- | --- | --- | --- | --- | --- | --- | --- | --- | --- | --- | --- |
|  |  | All<br>positives | Moderate<br>to strong<br>positives | All<br>positives | Moderate<br>to strong<br>positives | All<br>positives | Moderate<br>to strong<br>positives | All<br>positives | Moderate<br>to strong<br>positives | All<br>positives | Moderate<br>to strong<br>positives |
| DRG<br>neurons | IgG | 22<br>(71%) | 5<br>(16.1%) | 7<br>(58.3%) | 2<br>(16.7%) | 28<br>(36.4%) | 3<br>(3.9%) | 16<br>(37.2%) | 3<br>(7%) | 0.0054<br>(**) | 0.1193 |
| DRG<br>neurons | IgM | 28<br>(90.3%) | 10<br>(32.3%) | 12<br>(100%) | 2<br>(16.7%) | 21<br>(27.3%) | 3<br>(3.9%) | 14<br>(32.6%) | 0 | <0.0001<br>(***) | <0.0001<br>(***) |
| Schwann<br>cells | IgG | 3<br>(9.7%) | 2<br>(6.5%) | 0 | 0 | 6<br>(7.8%) | 0 | 3<br>(7%) | 0 | 0.7464 | 0.0348<br>(*) |
| Schwann<br>cells | IgM | 19<br>(61.3%) | 6<br>(19.4%) | 5<br>(41.7%) | 0 | 35<br>(45.5%) | 6<br>(7.8%) | 16<br>(37.2%) | 0 | 0.2256 | 0.0121<br>(*) |
| Any<br>reactivity | IgG/<br>IgM | 31<br>(100%) | 12<br>(38.7%) | 12<br>(100%) | 4<br>(33.3%) | 54<br>(70.1%) | 8<br>(10.4%) | 30<br>(69.8%) | 3<br>(7%) | 0.0009<br>(***) | 0.0004<br>(***) |

Overall, there was a greater chance of observing any anti-peripheral nerve cell IgG or IgM antibody reactivity in the ZIKV-GBS and ZIKV-OND patients’ sera, compared to non-complicated ZIKV (ZIKV-CON) and non-ZIKV control (CON) groups. These differences between groups were statistically significant (p=0.0009, Chi square test).

Additionally, we observed that IgG and IgM binding to DRG neurons or SCs were significantly more common in the ZIKV-GBS patients’ sera than in the ZIKV-CON group’s sera (p=0.0042, Fisher’s exact test); whereas there was no significant difference in the proportions of grouped anti-ZIKV sera and non-ZIKV sera (p>0.05, Fisher’s exact test) (see **Figure 1** for multiple comparisons).

**Figure 1.**
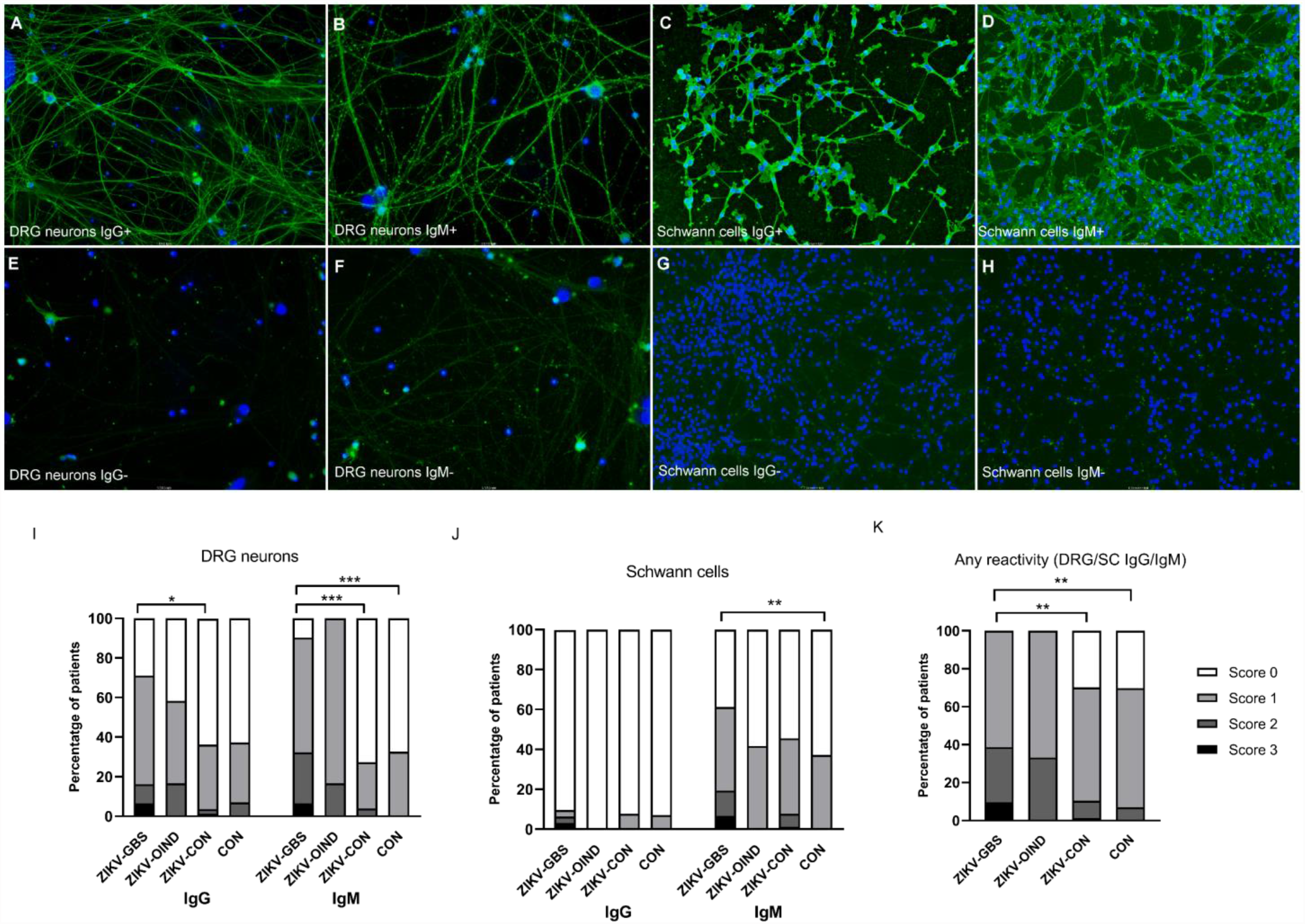
Reactivity of ZIKV patients’ serum IgG and IgM antibodies against primary cultured rat DRG neurons and Schwann cells. Immunofluorescent assay photomicrographs using ZIKV-GBS patients’ serum samples showing strong IgG (**A**) or IgM (**B**) antibody reactivity against DRG neurons, strong IgG (**C**) and IgM (**D**) antibody reactivity against Schwann cells compared to the negative control (CON) human sera which showed negligible/weak non-specific IgG (**E**) or IgM (**F**) antibody reactivity against DRG neurons or IgG (**G**) or IgM (**H**) antibody reactivity against Schwann cells. The percentage of IgG and IgM reactive antibodies of different intensities (0 = negative, 1 = weak, 2 = moderate and 3 = strong) from the different human patient groups (ZIKV-GBS, ZIKV-OND, ZIKV-CON and healthy controls (CON)) against DRG neurons, Schwann cells (SCs) or either DRG/SCs are shown in **I, J** and **K** respectively and the p-values of < 0.05, < 0.01 or < 0.001 are shown as *, **, and *** respectively.

### Screening against myelinating co-cultures

IgM binding to myelinating co-cultures was frequently detected with both the ZIKV-GBS patients’ and control group’s sera (**Table 2 and Supplementary Figure 2**). IgM reactivity was observed against the abaxonal membranes of myelinating SCs, axons, the processes of the non-myelinating SCs, as well as outpouchings of myelin (**Supplementary Figures 4A and B**). Overall, there was no significant difference in the proportions of ZIKV-GBS patients’ and control sera in terms of the frequency or pattern of positive IgM reactivity (p=0.3374, Fisher’ exact test) (see **Table 2** for multiple comparisons). IgM labelling of myelin outpouchings varied in intensity between the samples (**Supplementary Figures 4C and D**), however, immunofluorescence intensity at the myelin outpouchings normalised to the internode (**Supplementary Figures 4E and F**) was not significantly different between the ZIKV-GBS patients’, the ZIKV-CON patients’ or the healthy control group’s serum sample (One way ANOVA, F(2,106)=0.7417, p=0.4788) (**Supplementary Figure 4G**).

**Table 2.**
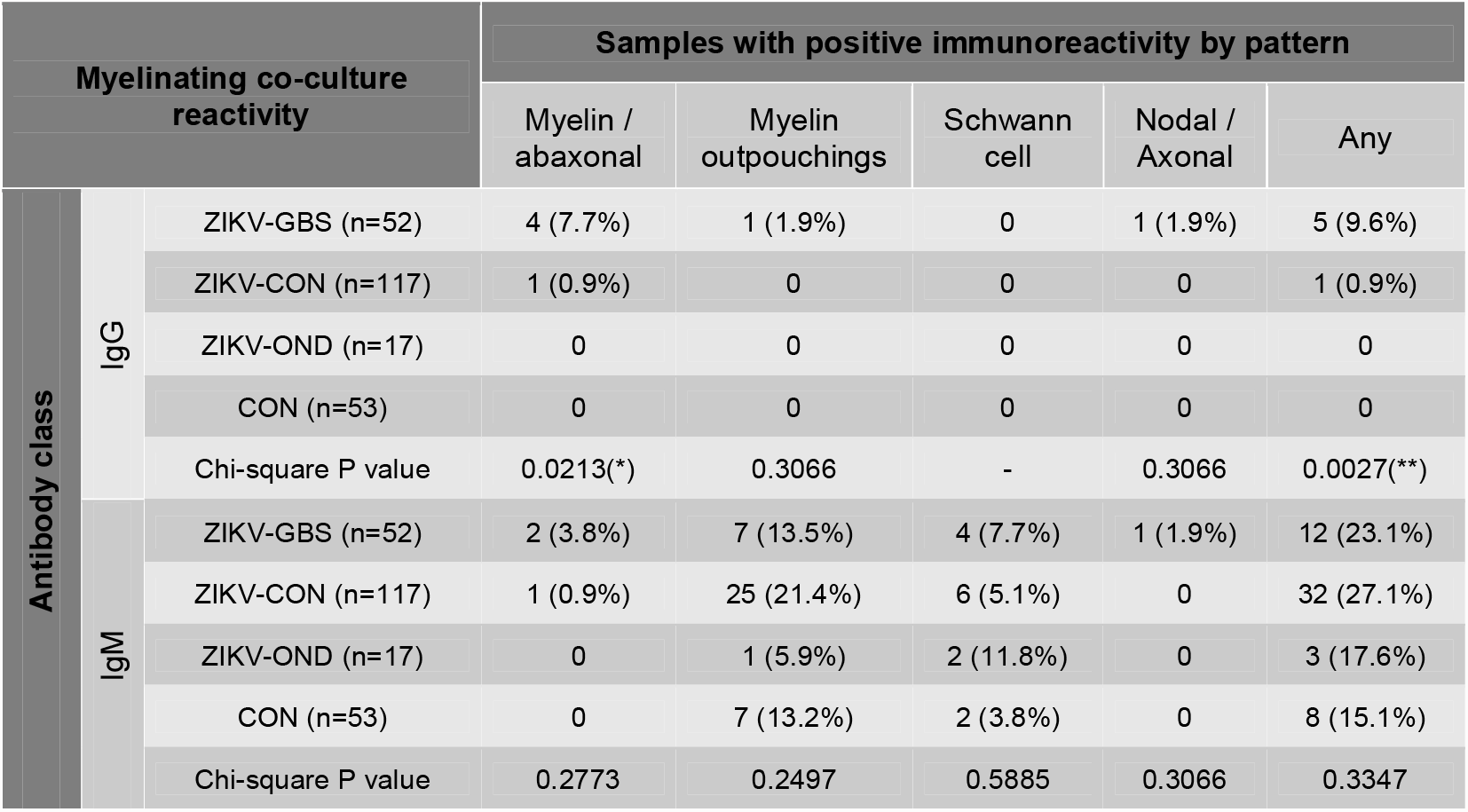
Frequency and statistical analysis of myelinating co-culture immunoreactivity.

| Myelinating co-culture reactivity |  |  | Samples with positive immunoreactivity by pattern |  |  |  |  |
| --- | --- | --- | --- | --- | --- | --- | --- |
|  |  |  | Myelin /<br>abaxonal | Myelin<br>outpouchings | Schwann<br>cell | Nodal /<br>Axonal | Any |
| Antibody class | IgG | ZIKV-GBS (n=52) | 4 (7.7%) | 1 (1.9%) | 0 | 1 (1.9%) | 5 (9.6%) |
|  |  | ZIKV-CON (n=117) | 1 (0.9%) | 0 | 0 | 0 | 1 (0.9%) |
|  |  | ZIKV-OND (n=17) | 0 | 0 | 0 | 0 | 0 |
|  |  | CON (n=53) | 0 | 0 | 0 | 0 | 0 |
|  |  | Chi-square P value | 0.0213(*) | 0.3066 | - | 0.3066 | 0.0027(**) |
|  | IgM | ZIKV-GBS (n=52) | 2 (3.8%) | 7 (13.5%) | 4 (7.7%) | 1 (1.9%) | 12 (23.1%) |
|  |  | ZIKV-CON (n=117) | 1 (0.9%) | 25 (21.4%) | 6 (5.1%) | 0 | 32 (27.1%) |
|  |  | ZIKV-OND (n=17) | 0 | 1 (5.9%) | 2 (11.8%) | 0 | 3 (17.6%) |
|  |  | CON (n=53) | 0 | 7 (13.2%) | 2 (3.8%) | 0 | 8 (15.1%) |
|  |  | Chi-square P value | 0.2773 | 0.2497 | 0.5885 | 0.3066 | 0.3347 |

IgG antibody reactivity was observed against different topographical domains within the co-cultures. Four out of 52 (7.7%) ZIKV-GBS patients’ sera contained IgG which bound the abaxonal membrane of a subset of myelinating SCs (**Table 2**), whilst showing no reactivity against non-myelinating SCs or axons (**Figures 2A and B**).

**Figure 2.**
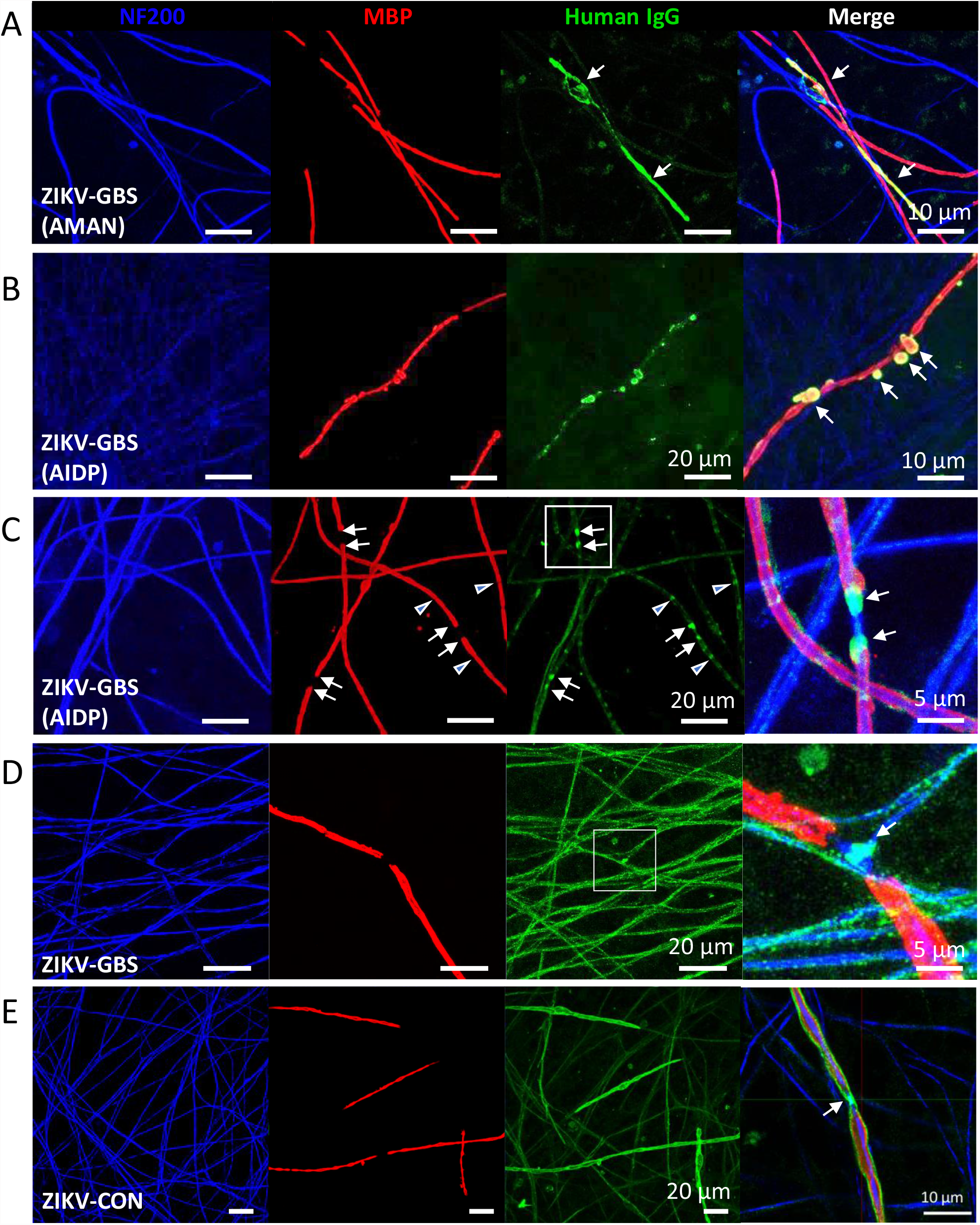
Patterns of ZIKV patients’ serum IgG reactivity against myelinating cell co-cultures. Examples of immunofluorescent photomicrographs of IgG binding patterns (green) of different ZIKV-GBS and ZIKV-CON patients’ sera in myelinated nerve cell cultures. (**A**) The selective labelling of individual internodes by IgG antibodies from one female ZIKV-GBS patient (*ZN060A*, 31-35 years old) with the AMAN subtype (a similar IgG binding pattern was seen in one other male ZIKV-GBS patient (*Cali006*, 41-45 years old). (**B**) Myelin outpouchings or ‘blebs’ labelled by IgG autoantibodies in the serum of a female ZIKV-GBS patient (patient D, *ZN005A*, 46-50 years old) with the AIDP subtype. (**C**) Myelin paranodes (arrows) and Schmidt-Lanterman incisors (arrow heads) labelled by IgG autoantibody reactions from the serum of a male ZIKV-GBS patient (*ZN039A*, 16-20 years old) with AIDP subtype. (**D**) Axonal and labelling with enrichment at the node shown by IgG autoantibody reactions from the serum of a male ZIKV-GBS patient (patient A, *Cali077*, 41-45 years old) fatal case. Nodal regions corresponding to insets in (C) and (D) are shown on the far right. (**E**) The labelling of the abaxonal membranes of all myelinating Schwann cells by the IgG autoantibodies in the serum of a female ZIKV-CON patient (patient B, *BQ008*, 41-45 years old) with previous uncomplicated ZIKV infection. Note that the staining is particularly intense at the Schwann cell microvilli overlying the node of Ranvier (right). Co-cultures are counterstained with NF200 (blue) and myelin basic protein (MBP) (red). Scale bars 5, 10 or 20 µm as indicated.

IgG from one ZIKV-GBS patient serum (male, 16-20 years old) was deposited on non-compact myelin at the paranodes and Schmidt-Lanterman incisures (**Figure 2C**). One ZIKV-GBS patient’s sera (patient A, male, 41-45 years old) labelled the axons of cultured neurons and their IgG antibody deposition was particularly concentrated at the nodal axolemma (**Figure 2D**). These patterns of IgG autoantibody reactivity were never seen with the control sera. The serum of one ZIKV-CON patient (patient B, female, 41-45 years old) also contained IgG antibodies, which gave an intense pattern of immunofluorescence over the abaxonal membranes of all myelinating SCs, and at the nodal microvilli (**Figure 2E**); no IgG labelling from patient B was present sensory neuron or SC monocultures (data not shown). Overall, IgG antibody reactivity against myelinating co-cultures was significantly more common in ZIKV-GBS patients’ sera (5/52; 9.6%) compared to all controls (1/187; 0.5%) (p=0.0021, Fisher’s exact test) (see **Table 2** for multiple comparisons).

### Assessment of ZIKV antibody-mediated axonopathy and demyelination

We next investigated the pathological potential of IgG autoantibodies in the two serum samples which reacted strongly against the cell co-cultures: patient A (**Figure 2D**) and patient B (**Figure 2E**). The axon and nodal binding of IgG antibodies present in the patient A’s serum was exclusively of the IgG1 subclass (**Supplementary Figure 5A**). The addition of patient A’s serum at a 1:100 dilution did not result in any axonal degeneration or other visible morphological nerve injury in the presence of complement in 20% NHS compared to that caused by a known complement-fixing anti-ganglioside IgM positive control antibody (**Supplementary Figure 5B**).

Antibodies reactive to myelinating SCs in patient B’s serum were also predominantly of the IgG1 subclass, with some IgG4 autoantibody subclass reaction against either SC processes or axons (**Supplementary Figure 6**).

The addition of patient B’s sera at a 1:100 dilution to co-cultures in the presence of 20% NHS resulted in complement deposition (C3c) around myelin internodes, and myelin fragmentation without apparent axonal degeneration (**Figures 3A, B**). Time-lapse imaging of fluoromyelin-stained internodes revealed retraction of the myelin from the node within hours of exposure to the anti-ZIKV serum in the presence of complement (**Figures 3C, D**). After 24h, multiple myelin internodes were either lost completely or significantly fragmented compared to baseline (0h) (**Figure 3E**). Anti-ZIKV serum treatment of the myelinating co-cultures in the presence of complement in the 20% NHS, but not anti-ZIKV serum or NHS alone, led to a significant loss of myelin coverage **(Figure 3F**), increased myelin fragmentation **(Figure 3G**) and shortening of internode length **(Figure 3H**), thereby confirming the effect seen using the time-lapse imaging (**Figure 3D**).

**Figure 3.**
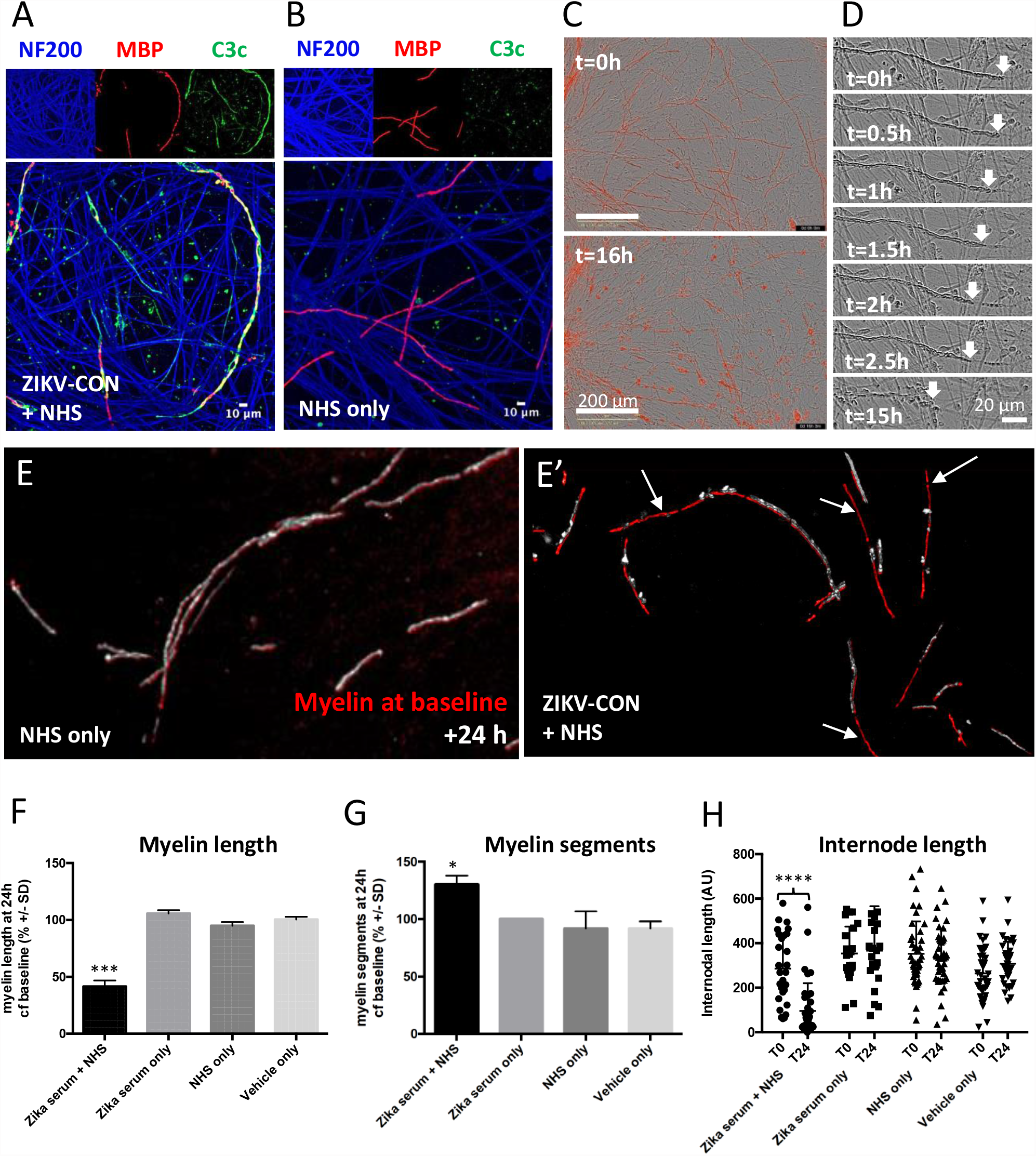
Complement-dependent demyelinating activity of ZIKV patients’ sera. The autoantibodies in the sera of ZIKV-CON patient B reacted with myelin internodes and fixed complement (C3c deposition; green) over myelin internodes (MBP; red) when complement was present from 20% normal human sera (NHS) (**A**) but not when only NHS was added to the cells (**B**). Counterstaining for NF200 (blue) and MBP (red) are also shown. (**C**) Fluoromyelin labelling (red) from live imaging of a myelinated co-culture before (*above*) and 16h after (*below*) incubation with patient B’s serum at a 1:50 dilution in the presence of complement in 20% NHS, while (**D**) shows the results of time-lapse stills of the receding myelin internode junctions shown from right to left over time (arrows). (**E/E’**) shows the overlaid example images of live myelin staining at baseline (red) and after 24h incubation (silver) with patient B’s serum with or without 20% containing complement. (**E’**) shows numerous myelin internodes are either lost completely or extensively fragmented after 24h (shown by arrows), while (**E**) without NHS containing complement the distribution of the myelin was essentially unchanged over 24h. (**F**) The total myelin length was reduced at 24h (**F,** *** p<0.001, one-way ANOVA with Bonferroni correction). (**G**) The number of myelin segments increased, (G, *p<0.05, one-way ANOVA with Bonferroni correction) in cultures treated with patient B’s serum with NHS containing complement compared to all control conditions (n=3 per group). (**H**) The mean internodal length was also significantly reduced at 24h compared to baseline in ZIKV-CON+NHS group only (**H**, ****p<000.1, 2-way ANOVA with Sidak’s correction).

### Antigen identification in myelinating co-cultures

The nodal/axonal binding ZIKV-GBS sera of patient A (**Figure 4A**) showed a similar binding pattern to sera from a separate study containing anti-CNTN1 antibodies (**Figure 4B**) [23]. Both sera maintained a similar IgG binding pattern when reacted with neuronal monocultures, thereby confirming the neuronal origin of both the unknown ZIKV-GBS auto-antigen (**Figure 4C**) and CNTN1 (**Figure 4D**). No immunolabelling was observed with healthy control serum (**Figure 4E**). In accordance with the nodal IgG labelling observed in the myelinating cell co-cultures, we tested for the nodal/paranodal candidate neurofascin (NF) in cell-based assays. Heterologous expression of NF155 and NF186 isoforms in a HEK cell-based assay confirmed their antigen-specific targeting by IgG1 antibodies in this ZIKV-GBS patient’s sera (**Figures 4F, G**) who had succumbed to lethal ZIKV-induced GBS. This ‘anti-pan-NF protein isoform’ IgG autoreactivity pattern was not detected in any of the other ZIKV-GBS patients’ or control sera.

**Figure 4.**
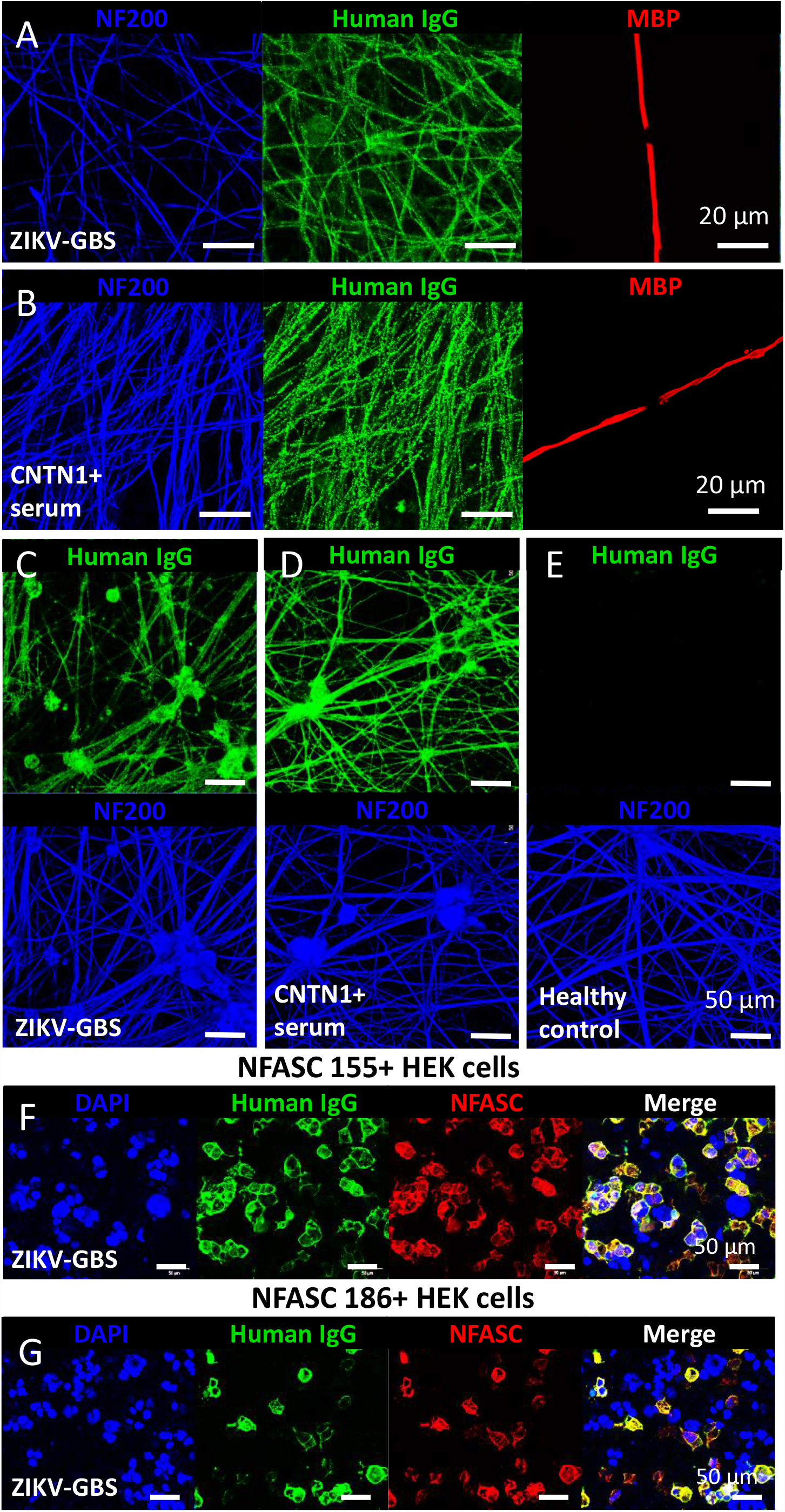
Transfected cell-based assays confirm pan-neurofascin reactivity of IgG antibodies within the serum of a patient with fatal, ZIKV-associated GBS. Immunofluorescent photomicrographs of (**A**) ZIKV-GBS patient A’s serum antibodies, reactive to unknown antigen enriched in axolemma and (**B**) a positive control patient’s serum with autoantibodies against the known nerve membrane antigen CNTN1 showed strong IgG (green) reactions against myelinated co-cultures, as well as in neuronal monocultures treated with (**C**) patients A’s serum, (**D**) the anti-CNTN1+ serum. (**E**) Healthy control serum was negative. Patient A’s IgG serum autoantibodies also reacted with HEK cells expressing the recombinant the NK155 (**F**) and NF186 (**G**) protein isoforms. Co-cultures are counterstained with NF200 (blue) and myelin basic protein (MBP) (red). Scale bars, 20 and 50 µm as shown.

### Immunoprecipitation

We employed a proteomic approach to identify candidate antigens from our live cell culture screening by isolating antigen-bound IgG antibodies from serum-treated cultures using protein G bead immunoprecipitation (IP) followed by high resolution C18 reversed-phase chromatography mass spectrometry (MS) analyses. We first validated this approach using patient A’s serum in neuronal cell monocultures. Three potential antigens were identified as enriched in the IP: NF, NCAM1, and NCAM2 (**Supplementary Data - Tab 1**), confirming that NF was a target antigen of patient A’s serum.

The abaxonal SCs binding ZIKV-CON patient B’s serum was compared with a patient with anti-NF155 antibodies by IP-MS, which showed a similar pattern of IgG binding (**Figures 5A, B, C and Supplementary Data – Tab 2**). A total of 28 proteins were significantly enriched by patient B’s sera including some complement components and seven membrane-related proteins: ANXA2, TGFBR3, NKCC1, NEP, CD44, ITGA6 and AHNAK2 (**Figure 5C**).

**Figure 5.**
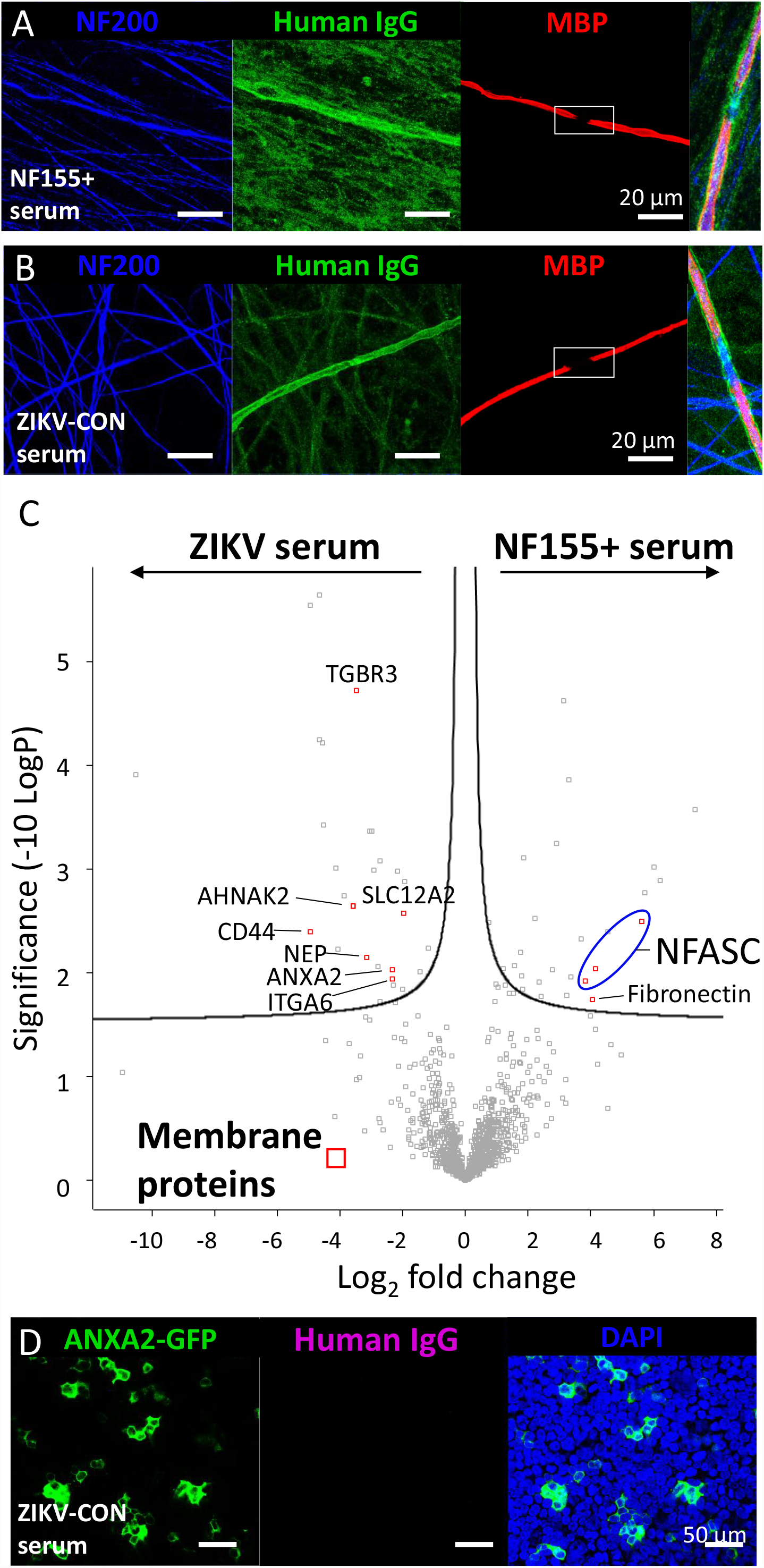
Identification of a ZIKV patient’s autoantibodies against myelin antigen: Immunoprecipitation and peptide mass spectrometry. Immunofluorescent photomicrographs of (**A**) a positive control patient’s serum with autoantibodies against the known nerve antigen NF155 and **(B**) ZIKV-CON patient B’s serum autoantibody reactions against unknown antigens enriched in abaxonal myelin, with higher magnifications of the nodal regions shown as insets (far right). Scale bars, 20 µm. Counterstaining for NF200 (blue) and MBP (red) are also shown. (**C**) Proteins identified in immunoprecipitates from ZIKV serum and NF155+ serum-treated co-culture lysates. t-test of enriched proteins presented as a volcano plot. iPSC lines from four donors were used to generate myelinating co-cultures, and immunoprecipitates were purified and analysed by mass spectrometry to give four biological replicates per serum sample. Significant data points were determined with a permutation-based false discovery rate (FDR) calculation (threshold = 0.1) and the significantly enriched membrane proteins are shown as red squares. NFASC (neurofascin) protein isoforms from a human and rat database are grouped together (blue circle). TGBR3, transforming growth factor receptor 3; AHNAK2, desmoyokin; SLC12A2, solute carrier 12/Na-K-2Cl cotransporter A2; NEP, neprilysin; CD44, P-glycoprotein 1; ITGA6, integrin alpha 6. The remaining peptides were identified as either serum components (immunoglobulin or complement proteins) or intracellular proteins. (**D**) ZIKV-CON patient B’s serum autoantibodies were however shown to be unreactive with the ANXA2 calcium binding membrane binding protein expressed in transfected HEK cells using immunofluorescent photomicrographic analysis.

Sera from two of the ZIKV-GBS patients (patient C and patient D) which showed moderate or strong reactivity against primary DRG neurons, and one from a healthy control, were also taken forward for IP-MS analysis. Antigen-bound IgG antibodies from patient C’s sera **(Supplementary Data - Tab 3)** and patient D’s sera (**Supplementary Data – Tab 4**), or secondary IgG anti-IgM bound to patient D’s IgM neurone autoreactive antibodies **(Supplementary Data – Tab 5**) were isolated from DRG neurons lysates by IP and processed for MS. In this study, six potential autoantigens (ALCAM, DPYSL2, NCAM1, CNTN1, L1CAM, VINC) were identified as enriched by these anti-ZIKV sera compared to the control sera.

### Evaluation of candidate antigens

Candidate antigens identified by IP-MS were evaluated by either ELISA or by heterologous expression in live cell-based assays. ELISA using human recombinant prosaposin and GM3 (alone or in combination), multiple human laminin isoforms and nidogen-1 failed to confirm these as targets of patient B’s serum IgG autoantibodies (**Supplementary Table 2**).

Heterologous expression in HEK cells failed to reveal binding of ZIKV-CON patient B’s serum IgG antibodies to ANXA2 (**Figure 5D**) or any of the large number of other candidate proteins in ELISA (**Supplementary Table 2**). Furthermore, neither ZIKV-GBS patient C’s or D’s serum IgG or IgM antibodies reacted against any of the candidate antigens by heterologous expression in these cell-based assays (**Supplementary Table 3**). We additionally analyzed IgG and IgM reactivity of anti-ZIKV sera against GAS6 and AXL proteins, which have been described as the receptors ZIKV virus entry to human neural cells as well as NrCAM, which shares 6 common peptide sequences with the ZIKV polyprotein. However, none of these sera reacted against these three candidate antigens or members of the AXL-GAS6 complex (**Supplementary Table 3**).

To account for the potential loss of antigen during the IP process, we subjected the lysates of myelinating cell co-cultures to electrophoresis on an acrylamide gel and after electro-blotting on nitrocellulose membranes the reactions of patient B’s sera autoantibodies were tested against these nerve cell antigens. Through this western blotting analysis, the patients’ IgG antibodies reacted strongly with two bands (**Supplementary Figure 7**). Manual filtering for membrane proteins associated with SCs or myelination identified three potential target antigens: PRX, ITGA7 and GFRA1 **(Supplementary Data – Tab 6**). However, no IgG reactivity to these particular antigens was observed using patient B’s serum on transiently-transfected cell-based assays (**Supplementary Table 2**).

### Chemical and enzymatic characterisation of target antigens

The intense labelling of the patient B’s serum in the lipid rich membrane and nodal microvilli of myelinating SCs (**Figure 6A**) led us to further investigate the target antigen characteristics using a biochemical approach. Solvent extraction of lipids from serum-treated and fixed myelinated co-cultures with mixture of chloroform, methanol and water as described previously [24] significantly reduced the amount of patient B’s serum IgG antibodies which bound to the abaxonal SCs membranes (**Figure 6B, C and D**), while the reactivity of commercial antibodies directed against NF2000 and MBP antigens were retained (**Figure 6B and C**). Solvent extraction did not affect labelling of human IgG to the GPI-linked protein CNTN1 (**Supplementary Figure 8**). Limited IgG antibody labeling remained at the paranodes (**Figure 6B**). Binding of patient B’s IgG antibodies was not however affected by prior neuraminidase treatment of the cell co-cultures (**Supplementary Figure 9**) compared with positive controls (**Supplementary Figure 10**) suggesting that the target antigen was not a sialylated glycolipid.

**Figure 6.**
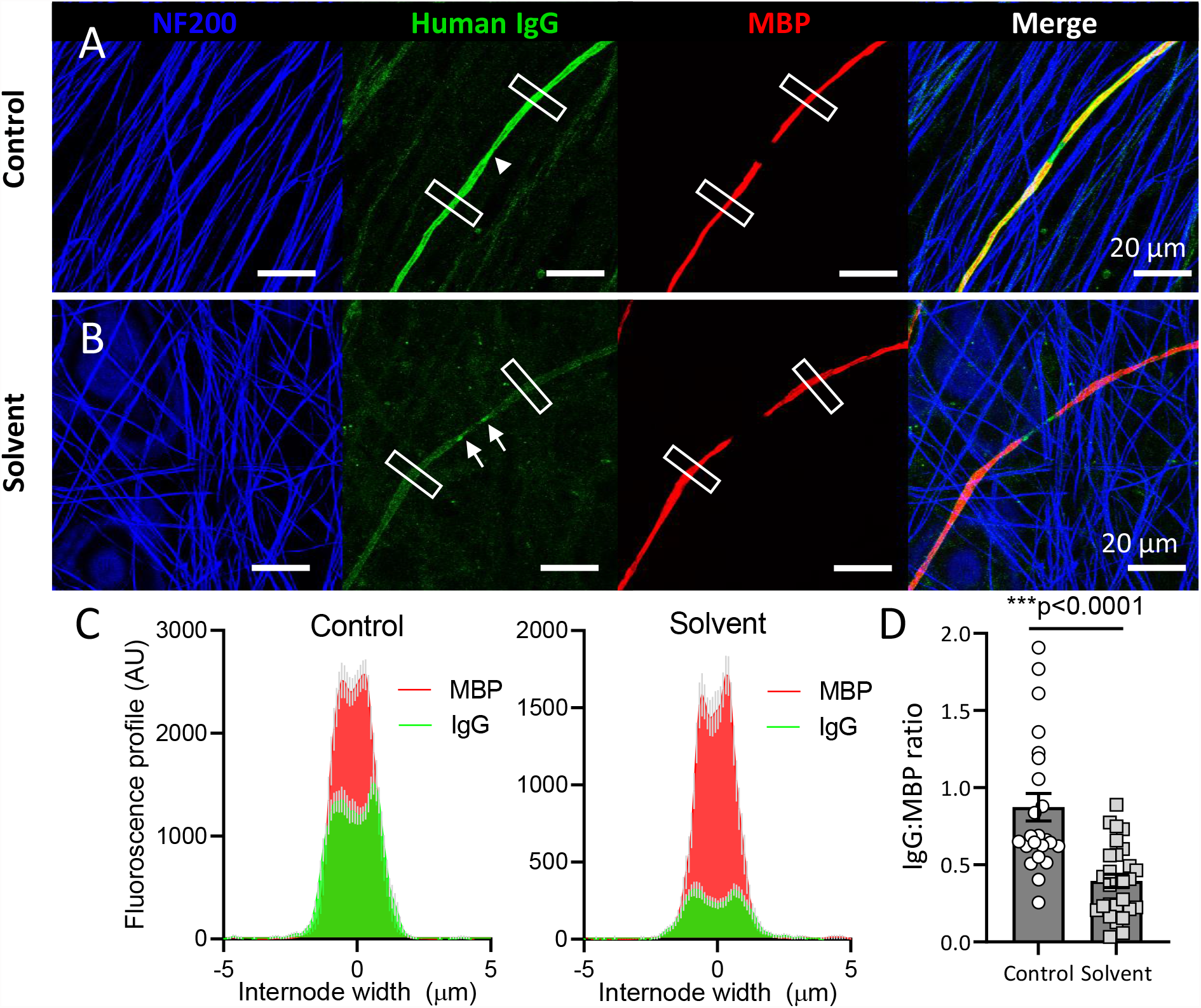
Removal of ZIKV-CON patient B’s serum IgG autoantibody target antigen(s) in myelinated sensory neuron cultures by lipid extraction. (**A**) Immunofluorescent photomicrographs of ZIKV-CON patient B’s serum IgG autoantibodies showing strong staining with untreated (Control) abaxonal membranes of all myelinating SCs, which was particularly intense at the Schwann cell microvilli overlying the node of Ranvier (arrowhead) while (**B**) very weakly staining these same cells after solvent treatment (Solvent). The control reactions for neuronal (NF200) and myelin (MBP) markers remained unchanged. For this study, the serum-treated fixed cultures were incubated with a mixture of chloroform, methanol and water at a ratio of 4:8:3 for 1h on ice to remove glycolipids. The white boxes indicate regions of fluorescence profiling per internode (5 x 20 µm). Arrows indicate weak paranodal IgG after lipid extraction. (**C**) The fluorescence intensity profiles of the human IgG autoantibodies and myelin basic protein (MBP) immunolabelling across the internodes of control and solvent-treated cultures. Data are presented as mean (± SEM in grey) of two internodes per field of view, 12-14 fields of view from two independent experiments. (**D**) The quantification of IgG:MBP fluorescence ratio. Student’s unpaired t-test, p<0.0001, t=5.053, n=24-28 internode profiles per group.

## DISCUSSION

Our study found that the humoral immune response in ZIKV-GBS cases was diverse and heterogeneous. We found neither a common antigen nor a common pattern of serum immunoreactivity against peripheral nerve structures despite all of these ZIKV-GBS patients sharing a common prodromal infection.

Consistent with a recent study of a modest cohort of ZIKV-associated GBS patients from Brazil,[11] we observed a small number of GBS patients with elevated anti-glycolipid IgG antibodies. However, we did not find a significant statistical association of ZIKV-GBS patient cases with any anti-glycolipid antibody signature, thereby supporting the results from a recent cohort study from Northeast Brazil. [25] The cases we did find may represent a subset of ZIKV induced GBS, or merely have been cases of classical onset GBS patients whose clinical course coincided with recent ZIKV infections.

Analysis of the ZIKV polyprotein showed that it contained peptide sequences which were also found in human proteins and that are known to be targets of inflammatory neuropathy autoantibodies.[26] However, while all ZIKV patient samples tested contained anti-ZIKV IgG antibodies, anti-nerve reactive autoantibodies showed a more discrete distribution, suggesting that any nerve-related antigenicity was not a consequence of humoral immunity to ZIKV antigens *per se*. Despite the high level of antigenic similarity between flaviviruses, we did not observe any specific nerve cell reactive autoantibodies in the DENV-infected control (DENV-CON) sera, in keeping with the rarity of GBS associated with recent DENV infections.[27] Although we observed one case of anti-pan-NF protein isotype IgG1 autoantibodies in our Colombian cohort (ZIKV-GBS patient A), prodromal infection is not a consistent feature of this rare and potentially fatal peripheral neuropathy subtype.[28]

By employing a comprehensive series of peripheral nerve cell culture models and antigen expression systems, we reveal a surprisingly heterogeneous autoantibody response in patients’ sera. Overall, IgG reactivity to rat SCs and the cell co-cultures was significantly higher, albeit infrequently, in the ZIKV-GBS patient group compared to all controls. IgM reactivity against rat DRG neurons and rat SCs was also significantly higher in ZIKV-GBS patient than in the control group. On the other hand, IgM immunoreactivity to myelin outpouchings, a feature reminiscent of the myelin foldings, swellings or tomaculae sometimes seen in certain hereditary neuropathies,[29] was remarkably common in both the ZIKV-GBS patient and control groups. The significance of these structures in the co-culture system is however unclear.

One patient (ZIKV-CON patient B), with an uncomplicated ZIKV infection, displayed striking IgG reactivity against abaxonal membrane of myelinating SCs that was particularly enriched at the nodal villi. These IgG1 subclass antibodies induced profound demyelination in our *in vitro* model system via a complement-dependent pathway, despite the patient lacking neurological symptoms. This apparent functional discrepancy might be accounted for by the use of rodent SCs in the co-culture system and a potential lack of cross-reactivity to human SCs, or by the target antigen(s) remaining hidden from circulating antibodies *in situ*.

By employing IP-MS in sera-treated myelinating co-cultures and DRG neurons, we revealed a number of candidate antigens related to either SC membranes, myelin formation, or neural cell adhesion. However, we could not confirm any candidate antigen reactivity by these patients IgG or IgM antibodies by either the ELISA or cell-based assays, reinforcing the challenges of antigen identification in the inflammatory neuropathies.[30] Nevertheless, the patterns of immunoreactivity specific to the patient groups, such as nodal, axonal and myelin internodes in the myelinating co-cultures, which were not observed in any control samples, suggest that the near-native antigen conformation in the cell co-culture system is a high-fidelity substrate for screening potentially pathogenic patients’ serum IgG antibodies.[19]

One of the limitations of our study is that our protocols are optimized for proteins and not for lipids. In fact, solvent-based lipid extraction significantly reduced reactivity of patient B’s IgG autoantibody reactions against myelinating SCs in the co-culture system, suggesting a lipidomic approach may be required. It is also possible that certain protein antigens were not detectable, either due to poor immunoprecipitation, or lack of representation in the peptide database used to compile our list of candidate antigens. An alternative approach is offered by high throughput antibody screening techniques such as the newly described Rapid Extracellular Antigen Profiling (REAP) technique,[31, 32] or chip-based proteome profiling [33], which has recently offered insights into the autoantibody repertoire of patients with chronic inflammatory demyelinating polyradiculoneuropathy (CIDP).[34]

Our findings mirror our recent screening of a separate cohort of 100 GBS patients, where we report a similarly heterogeneous reactivity of IgG and IgM to nerve-related antigens.[35] The narrow definition of the present GBS cohort, in which all patients associated with the same infectious insult, suggest that heterogeneity of the humoral immune responses in GBS may be a core feature of the disease and not necessarily due to heterogeneity of the patient cohorts.

## Data Availability

Mass spectrometry data are available via ProteomeXchange with identifier PXD028476. All other data produced in the present study are available upon reasonable request to the authors

http://www.proteomexchange.org/

## Acknowledgements

We thank Professors Peter Brophy and Sarosh Irani for kindly providing the periaxin antibody and healthy human control sera, respectively. We also thank Professor Robert MacLaren, Dr Tina Storm and Dr Ahmed Salman for their generous assistance with the IncuCyte time-lapse imaging. The authors would like to acknowledge the Department of Medicine at the Universitat Autònoma de Barcelona. CLl and LQ are members of the European Reference Network for Neuromuscular Diseases - Project ID No. 870177. pCSC-SP-PW-Nep (aka: pBOB-NEP) was a gift from Inder Verma. CD44S pBabe-puro was a gift from Bob Weinberg. anxA2-GFP was a gift from Volker Gerke & Ursula Rescher. ITGA6-bio-His was a gift from Gavin Wright. pcDNA3.1 Flag YFP hNKCC1 (NT17) was a gift from Biff Forbush. pBabe neo TGFBR3-HA was a gift from Kevin Janes.

---

**Supplementary Figure 1.**
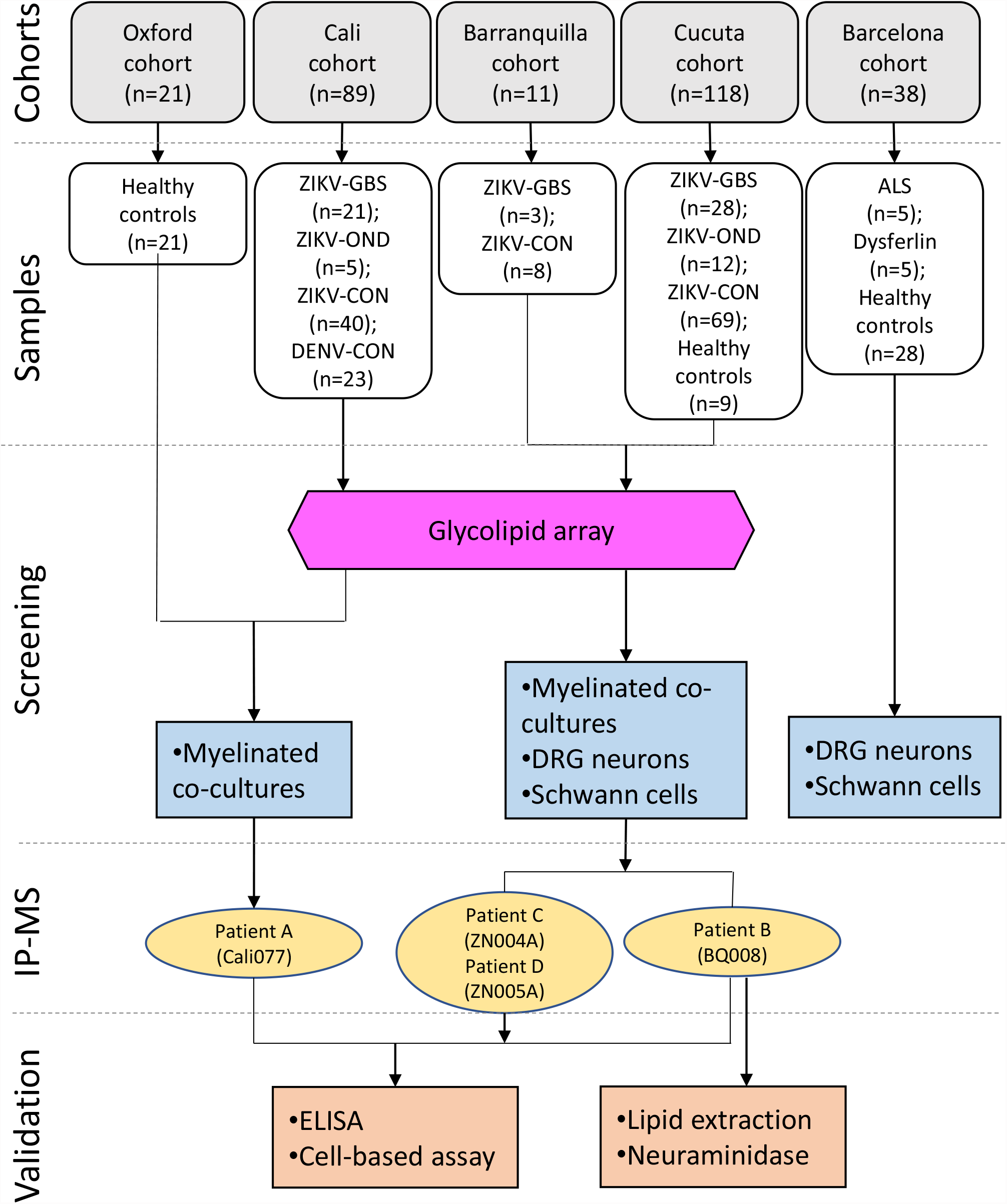
Diagram illustrating the patient cohorts and the experimental work flow with their serum samples. The numbers of patients’ serum samples collected in the different categories of ZIKV-GBS (ZIKV patients who developed GBS), ZIKV-OND (ZIKV patients who displayed other neurological diseases), ZIKV-CON (ZIKV patients who never developed clinical neurological disease), DENV-CON (DENV patients who never developed clinical neurological disease), ALS (patients with amyotrophic lateral sclerosis, a progressive nervous system disease that affects nerve cells in the brain and spinal cord, causing loss of muscle control), Dysferlin (patients with dysferlinopathy, a muscle diseases that have a slow progression of muscle weakness and atrophy) and healthy controls (n, number of patient samples). The patients’ serum samples were initially screened for their autoantibody reactions against different neuronal cell culture types and from which the sera from Patients A to D were tested further for their reactions in further assays.

**Supplementary Figure 2.**
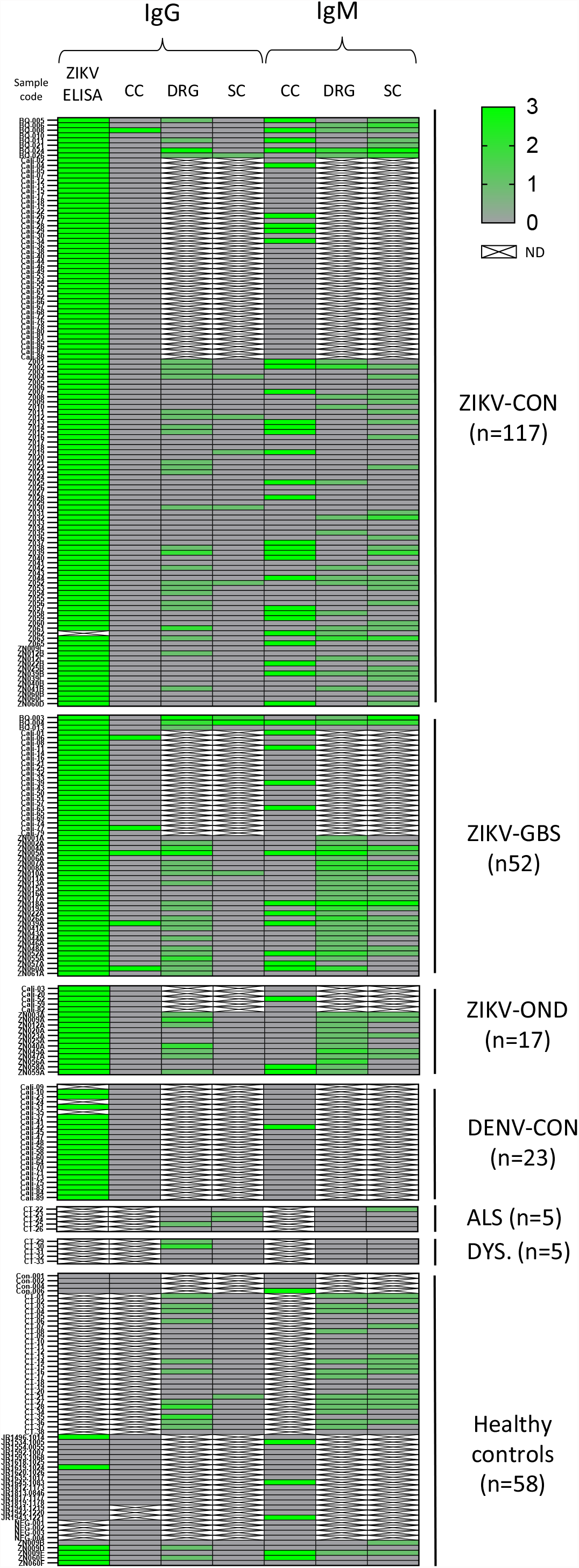
Simplified heat map showing all screening performed in ZIKV-GBS, ZIKV-OND, ZIKV-CON, DENV-CON, ALS, Dysferlin (DYS) and healthy controls (CON) patients. The reaction scores for each patients’ IgG and IgM antibodies from each group (ordered alphanumerically) against ZIKV (ELISA), rat dorsal-root ganglia neurons (DRG) and Schwann cells (SC) are shown by their colour intensity of each square (0=negative, 1=mild positive, 2=moderate positive, 3=strong positive), while their IgG and IgM antibody reactions with the myelinating cell co-cultures (CC) were scored as either negative (0) or positive (3). The serum samples that were not tested on a particular assay (due to lack of sample or availability) appear blank (ND, not determined).

**Supplementary Figure 3.**
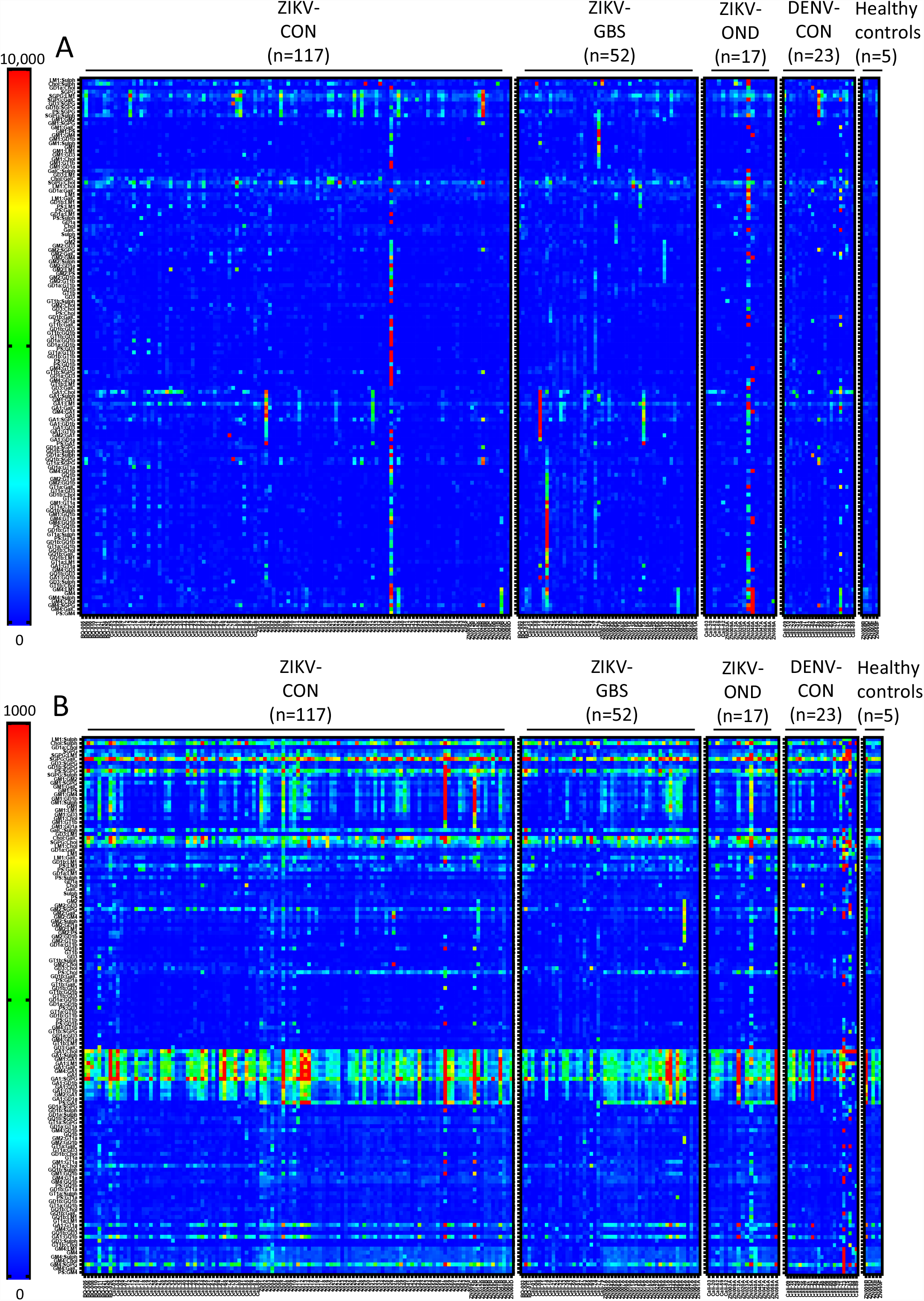
Glycolipid microarray results. Graphical display of the IgG (**A**) and IgM (**B**) antibody reaction intensities presented as heat maps from blue (negative) to red (strong) for each patients’ serum sample from each clinically classified group (ZIKV-GBS, ZIKV-OND, ZIKV-CON, CON) when reacted against 136 different glycolipid targets shown in the 136 rows. No significant differences were observed between the IgG or IgM antibody reactions against glycolipids and glycolipid complexes of the ZIKV-GBS patients compared with those of the control group (CON).

**Supplementary Figure 4.**
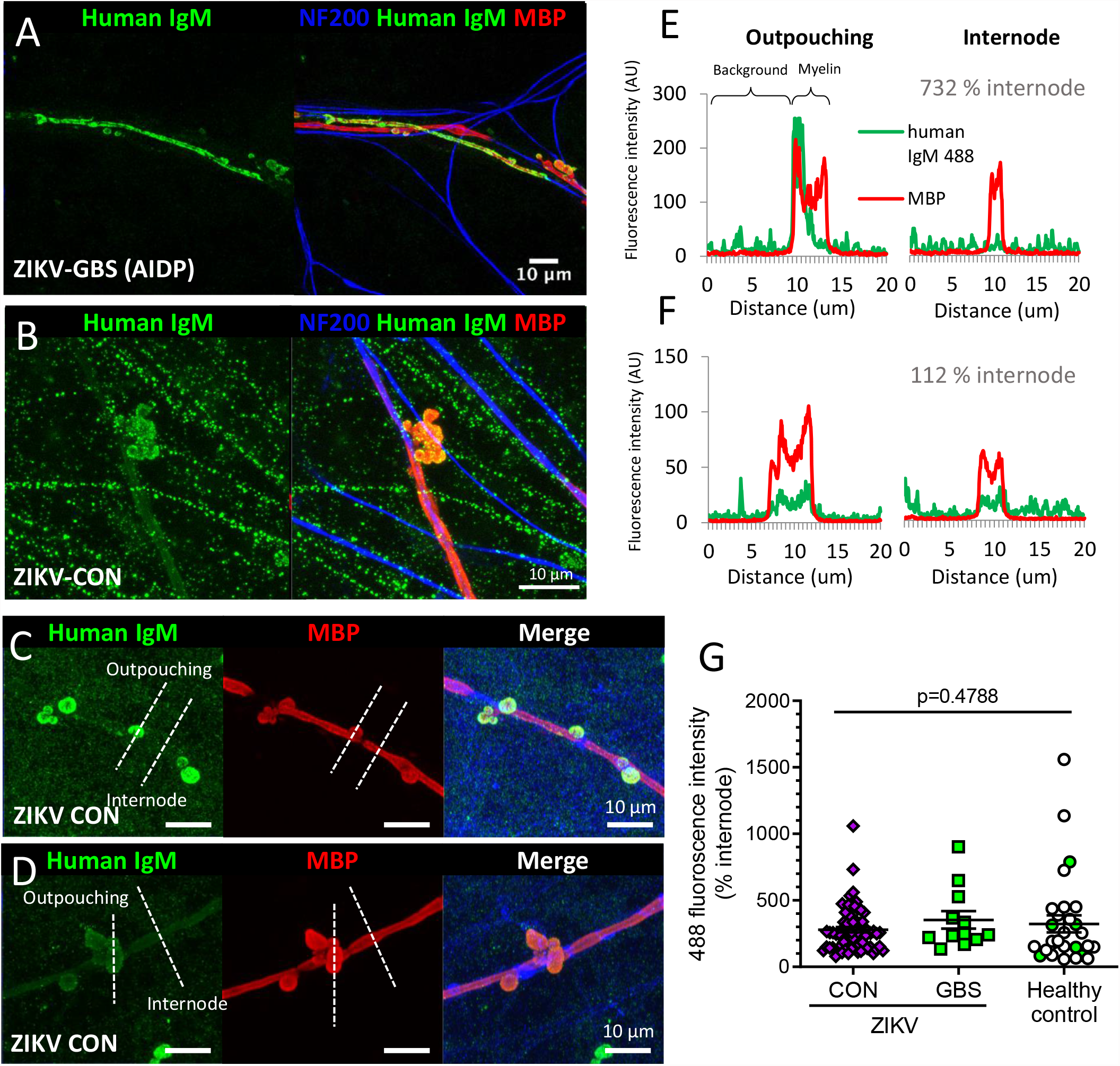
Patterns of ZIKV serum IgM reactivity against myelinating cell co-cultures. Immunofluorescent photomicrographs of IgM autoantibodies (**A**) from a ZIKV-GBS (AIDP) patient’s sera (patient D, *ZN005A*) with deposition on individual myelin internodes, (**B**) from a male ZIKV-CON patient’s serum (patient X, *ZN039B*, 41-45 years old) with deposition on non-myelinating Schwann cell processes, (**C**) from a female ZIKV-CON patient’s serum (patient Y, *Z040*, 51-55 years old) showing strong reactivity against myelin outpouchings and **D**) from another female ZIKV-CON patient’s serum (patient Z, *Z010*, 81-85 years old) showing weak reactivity against the myelin outpouchings. Counterstaining for NF200 (blue) and MBP (red) are also shown. Scale bars, 10 µm. The dashed lines illustrate the transection of myelin outpouchings and internodes for profile analysis of IgM signal intensity. (**E** and **F**) Fluorescent intensity profile plots of patients’ IgM autoantibodies (green) and myelin (red) across myelin outpouchings and internodes for the (**E**) strong and (**F**) weak labelling sera. (**G**) The IgM (Alexa 488) autoantibody fluorescence intensities of the ZIKV-GBS and ZIKV-CON patients sera compared to the healthy controls against myelin outpouchings normalised to internode labelling. One way ANOVA (all groups): F(2,106)=0.7417, p=0.4788). The healthy control serum samples which had detectable anti-ZIKV antibodies are represented by solid green circles.

**Supplementary Figure 5.**
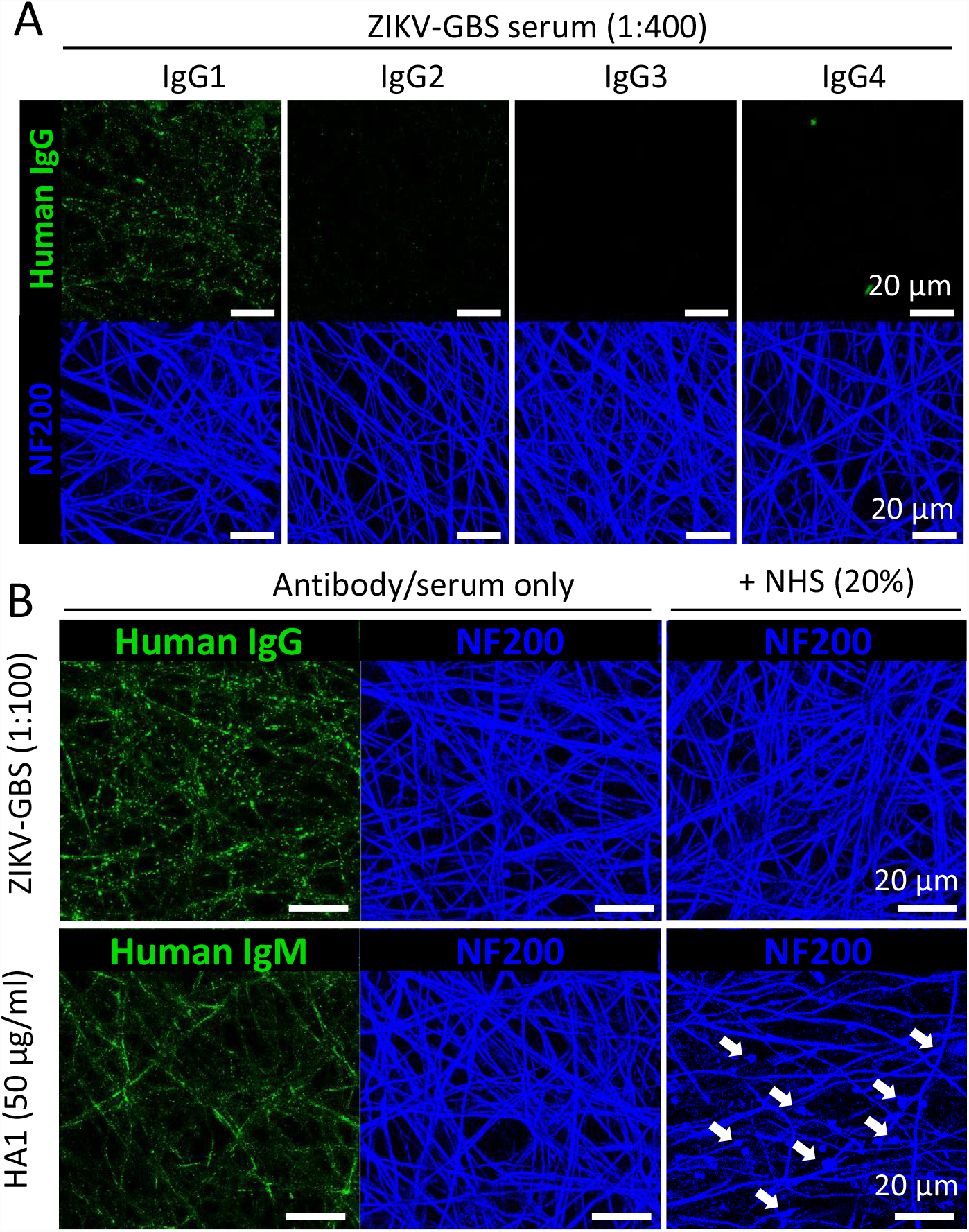
Axon-binding ZIKV-GBS serum antibodies are exclusively IgG1 subclass, but does not induce complement-mediated injury in neuronal culture. Immunofluorescent photomicrographs of (**A**) ZIKV-GBS patient A’s serum IgG autoantibodies reactions against myelinated sensory neuron cell co-cultures detected with IgG1, IgG2, IgG3 and IgG4 subclass-specific secondary antibodies and a subsequent fluorescent-labelled tertiary antibody (green) with NF200 labelling used as control counter-stain (blue). (**B**) Complement-induced injury to sensory neuron cell co-cultures. Incubation with ZIKV-GBS patient A’s serum at a 1:100 dilution or the anti-ganglioside antibody HA1 at 50 µg/ml for 1 hr followed by the addition of 20% normal human sera (NHS) containing complement overnight prior to fixation and immunolabelling. ZIKV-GBS patient A’s serum IgG autoantibody and HA1 binding were detected by fluorescent-labelled anti-human IgG and anti-human IgM secondary antibodies, respectively. Note the presence of axonal swellings (white arrows) and the reduced axon density in the HA1-treated cultures incubated with 20% NHS. No evidence of axonal injury was observed in the cultures pre-treated with the ZIKV-GBS patient A’s serum prior to NHS. Scale bars, 20 µm.

**Supplementary Figure 6.**
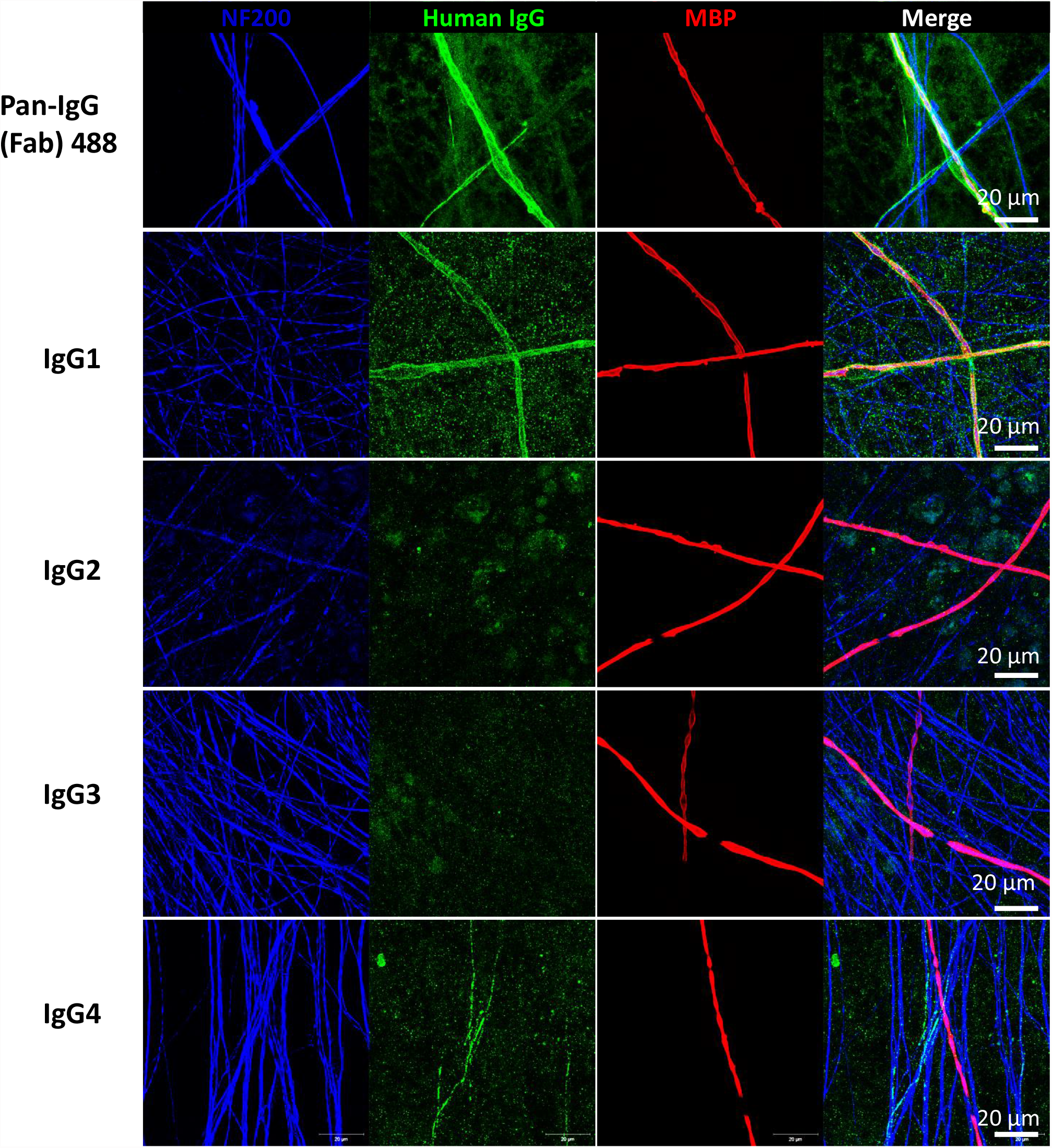
ZIKA-CON patient B’s sera autoantibodies predominantly of the IgG1 subclass. Immunofluorescent photomicrographs of the ZIKV-CON patient B’s serum autoantibodies against myelinating SC abaxonal-membranes and detection with either a pan-IgG subclass reactive or IgG1, IgG2, IgG3 and IgG4 subclass-specific secondary antibodies and a subsequent fluorescent-labelled tertiary antibody (green) showing their IgG autoantibodies predominantly of the IgG1 subclass but also with weaker IgG4 subclass antibodies. Counterstaining for NF200 (blue) and MBP (red) are also shown. Scale bars, 20 µm.

**Supplementary Figure 7.**
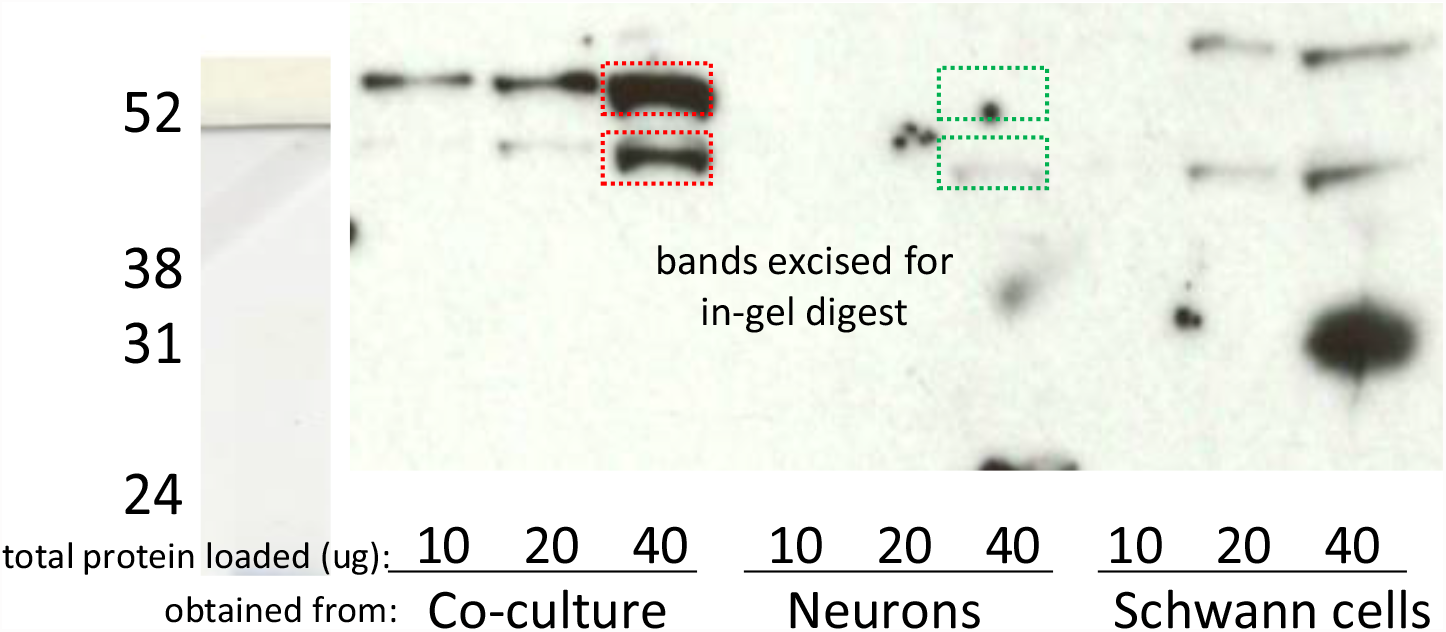
ZIKV-CON patient B’s IgG autoantibody reactions against nerve cell lysates in a western blotting assay. Different protein concentrations (10, 20 and 40 µg) of myelinated cell co-culture, sensory neuron and SC monoculture lysates were subjected to discontinuous acrylamide gel electrophoresis and then transferred onto nitrocellulose membranes. Western blotting with ZIKV-CON patient B’s serum was followed by sequential steps using peroxidase labelled anti-IgG secondary antibodies and ECL substrate and photographic detection. Two protein bands in the myelinated cell co-cultured lysate of relative molecular weights (Mr) of approximately 62 and 50 kDa were strongly detected by the serum IgG antibodies, but not in the neuronal or Swann cell lysates. The Mr standards of 52, 39, 31 and 24 kDa are shown in lane 1.

**Supplementary Figure 8.**
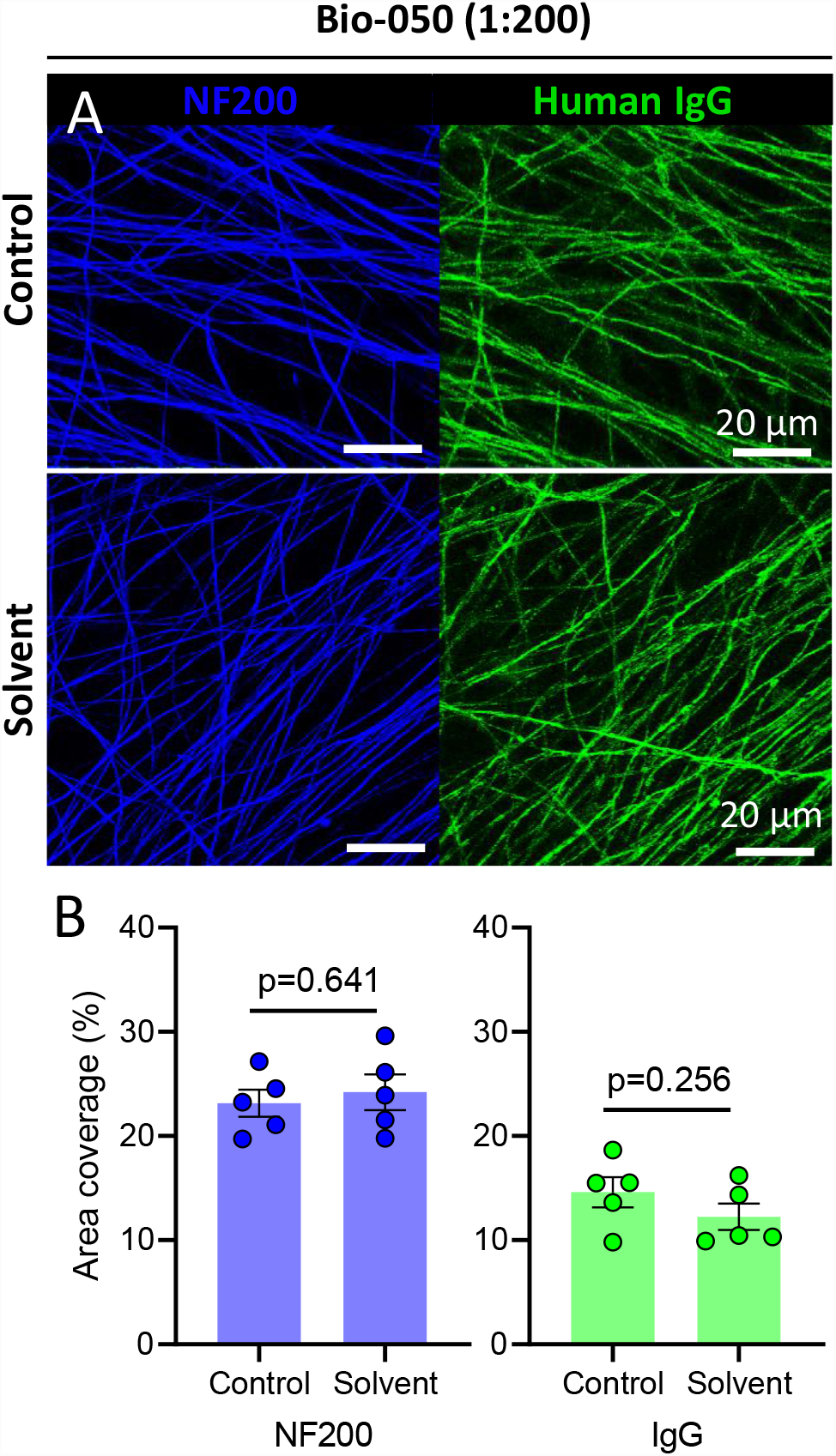
Human anti-contactin-1 (CNTN-1) serum IgG binding is not affected by lipid extraction. Immunofluorescent photomicrographs of the reactions of a diluted human serum which contained IgG antibodies against the GPI-linked protein CNTN1 added to myelinated sensory neuron cell co-cultures, followed either by (**A**) immediate (Control) immunolabelling for bound human IgG and neuronal (neurofilament, NF200) proteins or by solvent treatment (Solvent) prior to immunolabelling for bound human IgG and neuronal (neurofilament, NF200) proteins. Scale bars, 20 µm. (**B**) Quantification of the immunofluorescence labelling intensities for NF200 and human IgG antibodies per field of view were expressed as a percentage of the total area and showed that there was no significant loss of NF200 or human IgG-labelled immunofluorescence after before (Control) and after solvent treatment (Solvent). Identical acquisition and threshold settings were used for each field of view for each condition. The Student’s unpaired t-tests were used for these comparisons.

**Supplementary Figure 9.**
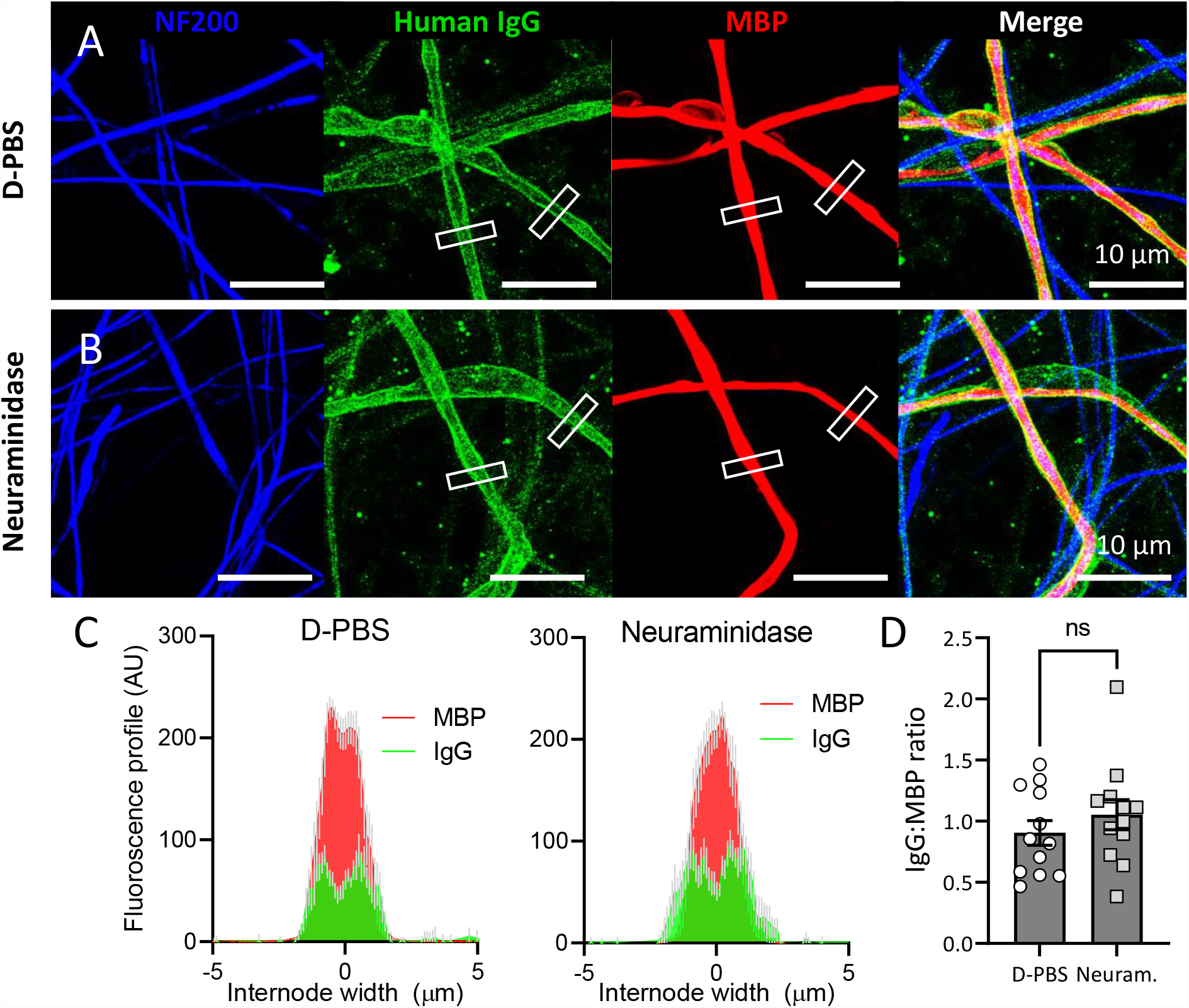
Effect of neuraminidase treatment of myelinating co-cultures on the reaction of ZIKV-CON patient B’s serum IgG autoantibodies. Immunofluorescent photomicrographs of live myelinating co-cultures treated with either (**A**) PBS containing divalent cations for 14 h (D-PBS) or (**B**) 1U/ml of neuraminidase for 14h (Neuraminidase) prior to addition of ZIKV-CON patient B’s serum IgG autoantibodies and the fluorescent labelled secondary antibodies or control labelling of NF200 and MBP. Scale bars, 10 µm. (**C**) The fluorescence intensity profiles (AU) of human IgG and MBP immunolabelling across internodes of control and solvent-treated cultures. The data are presented as the mean values (± SEM in grey) of two internodes per field of view, 6 fields of view from two independent experiments. (**D**) The quantification of IgG:MBP fluorescence ratios with the Student’s unpaired t-test, p=0.3605, t=0.9340, n=12 internode profiles per group.

**Supplementary Figure 10.**
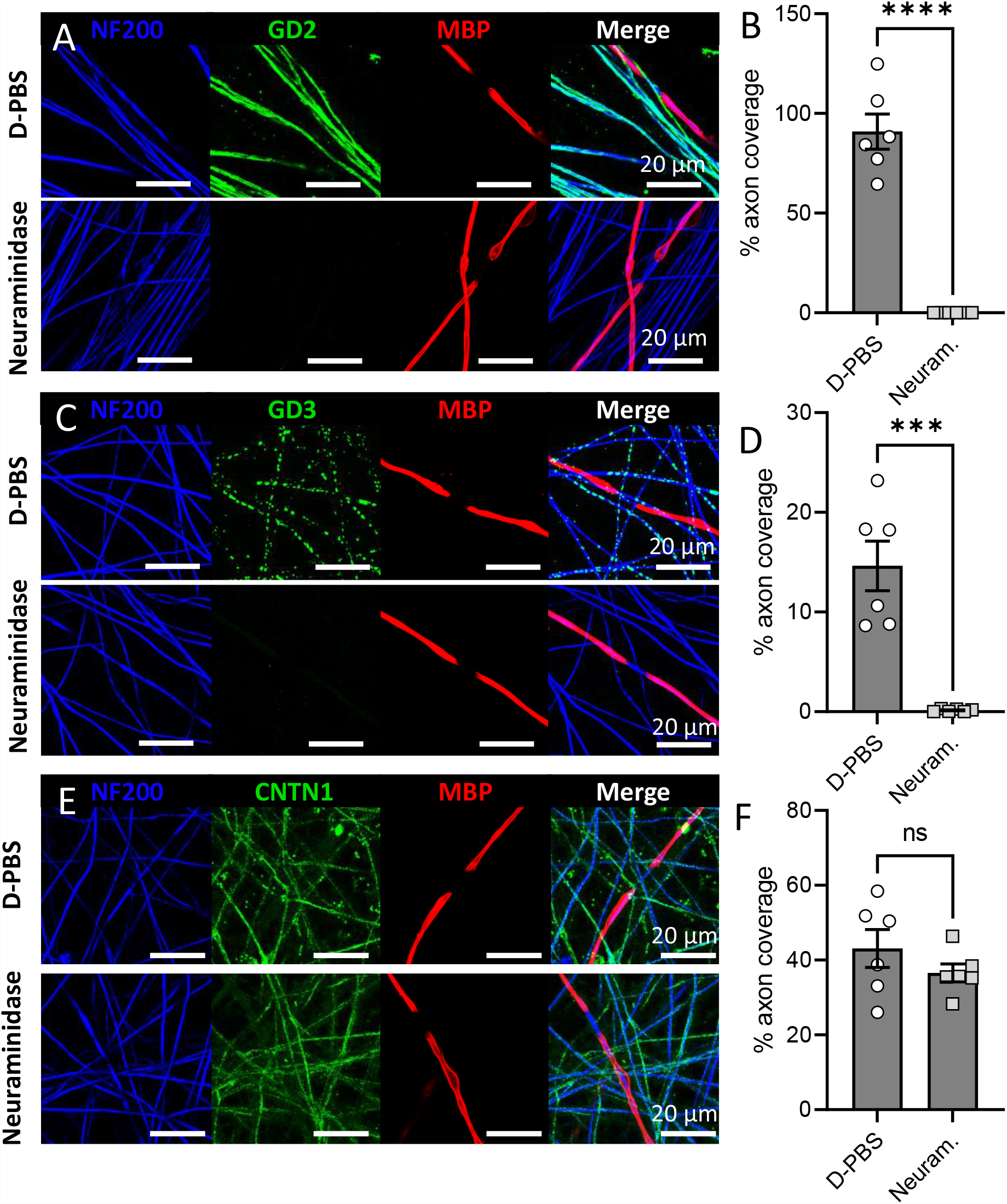
Sialic acid cleavage by neuraminidase treatment reduces binding of anti-disialosyl ganglioside but not anti-CNTN1 antibodies. Immunofluorescent photomicrographs of the reactions of (A) anti-GD2, (C) anti-GD3 anti-disialosyl ganglioside and (E) anti-CNTN1 GPI-linked protein antibodies (green) against live myelinating co-cultures after treatment with either 1U/ml of neuraminidase or D-PBS vehicle for 14 h. Counterstaining for NF200 (blue) and MBP (red) are also shown. Scale bars, 20 µm. The coverage of (B) anti-GD2, (D) anti-GD3 and (F) anti-CNTN1 IgG in the PBS and neuraminidase treated cultures expressed as a percentage of axon NF200 area.

