## Supplementary Methods and Tables for "Guillain-Barré syndrome following Zika virus infection is associated with a diverse spectrum of peripheral nerve reactive antibodies"

### SUPPLEMENTARY MATERIALS AND METHODS

Patient cohorts and sample acquisition

Electrodiagnostics

Rat dorsal root ganglia neurons and Schwann cells culture

DRG neuron and Schwann cell immunocytochemistry

Myelinating cell co-cultures

Myelinating cell co-culture immunofluorescence labelling

Chemical and enzymatic characterisation of target antigens

Assessment of serum-induced demyelination

Glycoarray

Immunoprecipitation and mass spectrometry

Protein electrophoresis and western blotting and target protein identification

ELISA

Transfected-cell based assays

Statistical analysis

### SUPPLEMENTARY TABLES

**Supplementary Table 1.** Summary of ZIKV patient cohorts and controls.

**Supplementary Table 2.** List of candidate antigens evaluated by ELISA.

**Supplementary Table 3.** List of candidate antigens for ZIKV-CON patient B's serum IgG autoantibody evaluated using transfected cell-based assays.

**Supplementary Table 4.** List of candidate antigens for patient C (IgG) and patient D (IgG/IgM) autoantibodies evaluated by transfected cell-based assays**.**

**Supplementary Table 5.** List of candidate antigens identified by immunoprecipitation and mass spectrometry, or literature review.

### SUPPLEMENTARY REFERENCES

### SUPPLEMENTARY MATERIALS AND METHODS

#### Patient cohorts and sample acquisition

Serum samples were collected from informed, consenting patients with ZIKV infections with and without neurological complications, patients with other infectious (e.g. DENV: dengue virus) and non-infectious diseases, as well as from healthy and convalescent ZIKV controls from three separate patient cohorts from different regions of Colombia: Cali (n=89), Cucuta (n=118) and Barranquilla (n=11). ZIKV patients with other inflammatory neurological diseases (ZIKV-OND) included myeloradiculopathy (n=1), encephalitis (n=5), meningoencephalitis (n=1), peripheral facial palsy (n=3) or transverse myelitis (n=7). The Spanish subjects (n=38) included 28 healthy controls and 10 patients with non-immune neuromuscular disorders amyotrophic lateral sclerosis (ALS) (n=5) and dysferlin muscular dystrophy (Dysferlin) (n=5). Sample collection was performed in compliance with Act 008430/1993 of the Ministry of Health of the Republic of Colombia, and the institutional review board of the Universidad del Rosario. Additional samples from healthy subjects with non-immune mediated neurological disorders, and nodal/paranodal antibody positive patients, were obtained for use from local collections in Oxford, UK (South Central - Oxford A Research Ethics Committee approval number 14/SC/0280) or Barcelona, Spain (The Sant Pau Biomedical Research Institute ethical approval number IIBSP-AUT-2016-69). Samples were coded and anonymized. Group sizes were based on sample availability and were not decided a priori. Whole blood was collected by venepuncture in SST II BD Vacutainer Tubes (BD Bioscience) and after clot retraction they were centrifuged at >2000 x g for 20-30 min. The separated serum was then aliquoted and stored at -80°C, with transportation on dry ice where required.

#### Electrodiagnostics

Electrodiagnostics was employed to classify GBS patients as either acute inflammatory demyelinating polyneuropathy (AIDP), acute motor axonal neuropathy (AMAN) or acute motor and sensory axonal neuropathy (AMSAN). Nerve conduction studies were evaluated according to Uncini's criteria (Uncini et al., 2018). Motor conduction studies, were evaluated in the median, ulnar, peroneal and tibial motor nerves whereas sensory nerve action potential and conduction velocity were measured in median, sural and ulnar nerves. F wave and H reflex were also registered. Where electrodiagnostics were unavailable patients were categorised as 'unclassified'.

#### Rat dorsal root ganglia neurons and Schwann cells culture

Animal procedures were performed according to Schedule 1 of the UK Home Office Animals (Scientific Procedures) Act 1986, and a protocol approved by the Animal Ethics' Committee of Hospital de la Santa Creu i Sant Pau. Dorsal root ganglia (DRG) were dissected from E16 rat embryos, dissociated and plated on glass coverslips coated with laminin (2.5 µg/ml) (Invitrogen, CA, USA) and poly-D-lysine (PDL) (25 µg/ml) (Sigma, MO, USA). These cells were then grown in neurobasal medium supplemented with 1x B27 and Glutamax (all Gibco, NY, USA) and mouse nerve growth factor (mNGF) (50 ng/ml) (Invitrogen). After 24 hours, cytosine arabinoside (Ara-C) (1 µM) (Sigma), floxuridine (40 µM) (Sigma) and uridine (40µM) (Sigma) were added to the medium to remove fibroblasts. This culture medium was replaced every other day until complete growth and differentiation of the DRG neurons was attained.

Schwann cells (SCs) were harvested from the sciatic nerves of 3 days-old rat pups, and cultured in Dulbecco's Modified Eagle Medium (DMEM) with high glucose (Thermo Fisher scientific, MA, USA), fetal bovine serum (FBS) (Gibco), mNGF and penicillin-streptomycin (Lonza, Basel, Switzerland). After 24h, Ara-C was added to the medium to remove fibroblasts. Three days later the medium was changed to a commercial Schwann cell medium (ScienCell, CA, USA) supplemented with FBS, penicillin-streptomycin, forskolin (Sigma) and neuregulin 1-β1 (NRG1-β1) (R&D systems). This medium was replaced every 2 days until the cells had sufficiently proliferated for harvest. These cells were dissociated by trypsin (0.25%, 5 min) diluted in DMEM and then plated on PDL coated coverslips until 70-80% confluence was reached. The cells were then fixed with 4% paraformaldehyde (PFA) in PBS, blocked for 1 hour with 5% goat serum in PBS and frozen at -80°C until immunocytochemistry experiments were performed.

#### DRG neuron and Schwann cell immunocytochemistry

Live DRG neurons were incubated for 1 hour with patients' sera diluted 1:100 (for IgG experiments) or 1:40 (for IgM experiments) in culture medium at 37°C. These cells were then fixed for 10 min with 4% PFA, followed by incubation with Alexa Fluor 488-conjugated goat anti-human IgG or IgM (1:1000 dilution) secondary antibodies (Thermo Fisher Scientific). Immunocytochemistry experiments on Schwann cells were performed on previously fixed and blocked frozen cells using patients' sera diluted 1:100 (for IgG labelling) or 1:40 (for IgM labeling) and appropriate secondary antibodies (1:1000 dilution). Coverslips were mounted with Vectashield with DAPI (Vector Laboratories, CA, USA) and the fluorescence signal intensities were scored by two independent researchers. Immunocytochemistry results were grouped according to three separate categories: moderate to strong positives (including scores 2 and 3), all positives (including scores 1, 2 and 3), and negatives (score 0). Images were obtained with an Olympus BX51 Fluorescence Microscope (Olympus Corporation, Tokyo, Japan).

**Myelinating cell co-cultures**

Myelinating cell co-cultures were prepared using human induced pluripotent stem cell (hiPSC)-derived sensory neurons and primary rat Schwann cells as previously described (Clark et al., 2017). hiPSCs from control subjects were obtained via the University of Oxford StemBANCC consortium. In brief, hiPSCs were differentiated into sensory neurons using a combination of small-molecule mediated dual SMAD (mothers against decapentaplegic family transcription factor) inhibition and Wnt pathway activation. On day 11 of differentiation, the sensory neuron precursors were seeded onto 13 mm diameter glass coverslips (approximately 20,000 cells per coverslip) or tissue culture-treated 10 cm petri dishes (approximately 200,000 per dish). Coverslips were previously cleaned by acid-washing, and coated with PDL (10 µg/ml) and reduced growth-factor Matrigel (Corning). Neurons were matured for 2-4 weeks in neurobasal media supplemented with 1x N2 (Cat. 17502-048), B27 (Cat 12587-010), Glutamax (Cat. 35050-038) and 1x antibiotic-antimycotic (penicillin, streptomycin and amphotericin) mixture (Cat. 15240-062) (all Gibco, Life Technologies) ('complete' neurobasal medium) plus recombinant human β-NGF (rhNGF) (Cat. 450-01, Peprotech), NT3 (Cat. 450-03, Peprotech), GDNF (Cat. 450-10, Peprotech), and BDNF (Cat. PHC7074, Life Technologies). All growth factors were used at 25 ng/ml. Primary Schwann cells were isolated from rat pups (P2-3) according to the protocol above, added to the neuronal cultures (25,000 cells per coverslip; 200,000 cell per 10 cm dish) and allowed to proliferate and align with the axons for 1 week in basal media containing insulin (5 µg/ml) (Sigma), holo-transferrin (100 µg/ml) (Sigma), rhNGF (25 ng/ml) (Peprotech) (Sigma), selenium (25 ng/ml) (Sigma), thyroxine (25 ng/ml) (Sigma), progesterone (30 ng/ml) (Sigma), triiodothyronine (25 ng/ml) (Sigma) and putrescine 8 µg/ml (Sigma) in DMEM/F12 media (Gibco, Life Technologies). From this point on, these cell cultures were maintained in 'myelination medium' containing 5% CS-FBS, ascorbic acid (25 µg/ml), phenol-free Matrigel (1:300 dilution) (Corning) and hrNGF (25 ng/ml) in 'complete' neurobasal medium. Myelinating cultures were matured for at least 4 weeks before use in subsequent experiments.

**Myelinating cell co-culture immunofluorescence labelling**

To assess the immuno-reactivity against myelinating co-cultures, sera from ZIKV-exposed subjects was diluted 1:100 in ‘complete’ neurobasal media supplemented with rhNGF (25 ng/ml) bovine serum albumin (BSA, 1%) and added to these live co-cultures for 1h at 37°C. These cultures were then washed in three changes of DMEM containing 20mM HEPES (DMEM/HEPES) and fixed in 1-2% paraformaldehyde (PFA) for 30 minutes at room temperature (RT). Alexa Fluor-488 conjugated goat anti-human IgG (H+L) (A11013, Life Technologies), F(ab’)2 goat anti-human IgG Fc (H10120, Life Technologies) or goat anti-human IgM (μ chain specific) (A21215, Life Technologies) secondary antibodies were then added at a 1:750 dilution for 1h RT in DMEM/HEPES + 1% BSA, before washing in DMEM/HEPES. For IgG subclass determinations, mouse anti-human IgG 1 to 4 (I2513, I5635, I7260, I7385, Sigma) were added at a 1:50 dilution in DMEM/HEPES plus 1% BSA for 1h at RT, followed by goat anti-mouse IgG Alexa 488 (1:1000) (A-11029, Life Technologies).

For the assessment of complement deposition, rabbit anti-human complement C3c (A006202-2, Dako) at a dilution of 1:200 was added and after reaction and washing the bound anti-C3c IgG was detected by Alexa 488-conjugated goat anti-rabbit IgG (A-11008, Life Technologies) (1:1000 dilution). These cells were then permeabilised with ice-cold 100% methanol for 25 minutes, rinsed in PBS, blocked in 5% NGS for 1h at RT and incubated with chicken anti-neurofilament heavy chain (NF200) (ab4680, Abcam) (1:10,000 dilution) and rat anti-myelin basic protein (MBP) (ab7349, Abcam) (1:500 dilution) primary antibodies overnight at 4°C to label neuronal processes and myelin internodes, respectively. After washing in PBS, these cells were incubated with biotinylated goat anti-chicken IgY (BA-9010, Vector) (1:500 dilution) and Alexa 546-labeled goat anti-rat IgG (A11081, Life Technologies) (1:1000 dilution) secondary antibodies for 1h at RT, followed by streptavidin-conjugated Pacific Blue (S11222, Life Technologies) (1:500 dilution) for 45-60 min at RT. The coverslips were then mounted onto microscope slides (Superfrost Plus, Thermo Scientific) using Vectashield mounting medium (H1000, Vector Laboratories) and stored at -20°C until imaging. The images were then acquired using confocal laser scanning microscopy (LSM 700, Zeiss) with a 63x oil-immersion lens (with a 10x eye-piece = 630x magnification) and exported as maximum intensity projection of 4-5 z-sections at 1 µm intervals. These images were then assessed for IgG or IgM reactivity to distinct topographical domains by an operator blinded to the diagnostic category of the samples. These serum reactions were categorised as either positive or negative, and the topographical binding patterns were recorded for the following categories: myelin/abaxonal (including outpouchings), Schwann cell and nodal/axonal.

#### Chemical and enzymatic characterisation of target antigens

For lipid-extraction, serum-incubated cell cultures were washed with PBS and fixed with 4% PFA for 30 min at RT before treatment with or without a solvent mixture of chloroform, methanol and water at a ratio of 4:8:3 (Svennerholm and Fredman, 1980) for 1h on ice. For the enzymatic digestion of sialylated antigens, live myelinated cell co-cultures were pre-treated with 1U/ml neuraminidase (from Clostridium perfringens, Sigma, N2876) in PBS containing $Ca^{2+}$ and $Mg^{2+}$ (D-PBS) for 14 h at 37°C. After washing 3 times in D-PBS, these cultures were subsequently incubated with patients' serum for 1h at 37°C before washing in PBS and fixing with 2% PFA for 20 min at RT. Subsequently, the coverslips were immunolabelled for human IgG, MBP and NF200 following the protocol above.

#### Assessment of serum-induced demyelination

Myelinating cell cultures were incubated for 1h in 'complete' neurobasal media supplemented with fluoromyelin red (F34652, Fisher Scientific) (1:300 dilution), washed 3 times in PBS, and placed back into myelination medium. Baseline myelin coverage was imaged on an inverted microscope (Leica DM IL) fitted with LED fluorescence excitation (pE-300, CoolLED) and the results were obtained using a digital camera. Serum-free myelination medium was then supplemented with or without ZIKV patients' sera at a 1:100 dilution containing 20% normal human serum (NHS) as a source of serum complement proteins and added to these co-cultures. After 24h incubation at 37°C, the culture medium was replaced, re-stained with fluoromyelin and re-imaged. To illustrate the change in myelination after the 24h incubation period with serum compared to baseline, the layers images were overlaid using the transparency function in Photoshop (Adobe). For quantification, the myelin internodes on the original, unprocessed baseline and 24h images were counted and measured using the line measurement function on ImageJ (NIH). Time-lapse images of demyelination during ZIKV patients' serum-treatment (1:50 dilution) at 37°C were acquired every 15 min at 10 times magnification using phase contrast and red channel (565–605 nm excitation) epi-fluorescence on an IncuCyte S3 live-cell imager (Sartorius). Fluoromyelin staining was used to confirm myelin internodes at the beginning (0h) and end (16h) of the experiment. The complement-fixing human monoclonal antiganglioside IgM antibody HA1 used a positive control was cloned after EBV transformation of peripheral blood mononuclear cells taken from a patient with CANOMAD, as previously described (Willison et al., 1996).

**Glycoarray**

Serum samples were screened on a glycolipid microarray as previously described (Halstead et al., 2016). In brief, all sera were screened against a panel of 16 single glycolipids (GM1, GM2, phosphatidylserine, GM4, GA1, GD1a, GD1b, GT1a, GT1b, GQ1b, GD3, SGPG, LM1, cholesterol, GalC and sulphatide) and 120 heteromeric 1:1 (v:v) complexes, printed in duplicate. Array slides were blocked with 2% BSA in PBS for 1 hour at room temperature, prior to addition of the patients' serum samples.

FAST incubation chambers and frames (GVS, USA) were used to separate the individual arrays on each slide, to which 100µl of each human serum sample, diluted at 1:50 in 1% BSA/PBS was added for 1h at 4°C. Following two 15 min washes in 1% BSA/PBS, anti-glycolipid antibody reactivities were detected using fluorescently conjugated, isotype specific, anti-human IgG and IgM antibodies (see above). Arrays were scanned with a Genepix 4300A (Molecular Devices, USA) and the average of duplicate median fluorescent intensity for each glycolipid target was calculated.  Anti-glycolipid antibody binding intensities were displayed on a heat map (MeV software) using Pearson's correlation hierarchical clustering.

**Immunoprecipitation and mass spectrometry**

Human sera that showed moderate or strong (scores 2 or 3) reactivity against rat DRG neurons were used for the immunoprecipitation (IP) experiments using the same cell types as substrates as previously reported (Siles et al., 2018). Briefly, protein A and G agarose beads (Invitrogen) or anti-human IgM-agarose antibody (Sigma) were used to isolate human serum IgG or IgM which was allowed to bind overnight to the rat DRG neuron cell culture extract. The precipitated proteins were detached from the agarose beads using Laemmli sample buffer (BioRad, CA, USA) containing with 5% 2-mercaptoethanol and separated by electrophoresis. The protein bands which appeared using the patients' serum IPs but not in the controls, were analysed by mass spectrometry. Proteins were selected as candidate antigens using Anaxomics (Anaxomics Biotech SL, Spain). Before filtering, the software deleted non-specific and low-signal antigens from the uploaded data, and only entries that met the following criteria were analysed: a) protein scores >100, b) peptide sequence coverage >5% or c) two or more peptides identified with the absence of the same criteria in the control sample. After this treatment, the software applied a set of sequential filters of inclusion, including: nerve or brain expressed proteins, membrane proteins and surface proteins.

Correspondingly, myelinating cell co-cultures or sensory neuron monocultures were scaled up into 10 cm petri dishes to provide a substrate for IP of the positive human sera identified in the co-culture system. After >8 weeks of maturation in myelination medium, the sera from selected ZIKV-exposed and control subjects were diluted 1:100 in neurobasal/NGF/1% BSA and added to live co-cultures for 1h at 37°C. The cells were then washed 3 times with ice-cold PBS and lysed for 15 minutes on ice with 500 μl of RIPA buffer supplemented with a 1X protease inhibitor cocktail (Halt, ThermoFisher Scientific). These cells were then gently scraped from the plate into a 1.5 ml tube and incubated for 30 min at 4ºC with gentle mixing / inversion. Homogenisation was ensured with repeat pipetting and samples then centrifuged in a benchtop centrifuge for >10,000 x g for 5 min at 4°C. The supernatants were collected and incubated with 50 μl of protein G dynabeads (Invitrogen) for 2hr at 4ºC on a rotating mixer and then washed 3 times with 200 μl PBS using a magnetic rack. Elution was performed using 200 µl of 0.1M glycine/HCl (pH 2.6) added for 2 min at RT and repeated a further 2 times (total 600 µl).

For mass spectrometry, IP eluates from myelinating cell co-cultures were prepared by chloroform:methanol precipitation and in-solution trypsin digestion. These peptides were then subjected to C18 reversed-phase HPLC column chromatography and analysed by data-dependant MS/MS on a ThermoFisher Fusion Lumos mass spectrometer. Antigen-bound IgG from the ZIKV patients' serum was compared with serum containing IgG reactive with known nerve cell antigens to enable calculations of their relative protein enrichments. For comparison of the ZIKV-CON patient B's serum and the anti-neurofascin-155 (NF155) antibody positive sera, IPs were generated from lysates from myelinated cultures of sensory neurons derived from four donor hiPSC lines. For statistical analysis of biological replicates, sample peptide data were aligned with concatenated human, rat and ZIKV reference proteomes (UniProt.org) and split using Progenesis QI (Nonlinear Dynamics) to generate raw abundance values. Protein enrichment was quantified by transforming their normalised abundance values and presented in a volcano plot using Perseus (MaxQuant) (Tyanova et al., 2016). For single sample comparison of ZIKV-GBS patient A, label free quantification was performed using Peaks software v7 (Bioinformatics Solutions Inc.). For in-gel digests, raw files from each LC-MS/MS injection were converted to a MASCOT Generic Format (MGF) and searched on the MASCOT server (version 2.5.1) against the UPR Homo sapiens and Rattus Norvegicus databases (UniProt.org). Unique peptides were exported and compared for presence or absence in either co-culture or neuronal culture lysates. Full data sets are available in the **Supplementary Data** file. The mass spectrometry proteomics data have been deposited to the ProteomeXchange Consortium via the PRIDE (Perez-Riverol et al., 2019) partner repository with the dataset identifier PXD028476 and 10.6019/PXD028476

#### Protein electrophoresis and western blotting and target protein identification

Lysates from mature myelinating cell co-cultures, hiPSC-derived sensory neurons monocultures, and primary rat Schwann cells were prepared with 500μl RIPA buffer supplemented with a 1X protease inhibitor cocktail (Halt, ThermoFisher Scientific). Total protein content was quantified using a bicinchoninic acid assay kit (Pierce BCA Protein Assay Kit, ThermoFisher Scientific) according to manufacturer's instructions. 10, 20 and 40μg of each lysate was prepared in lithium dodecyl sulfate (LDS) sample buffer, heated to 37°C for 10 minutes, loaded into separate wells of 4-12% continuous acrylamide Bis-Tris mini-gels and separated by electrophoresis in MOPS running buffer (ThermoFisher Scientific). The separated proteins were then either wet transferred to 0.45 μm pore-sized nitrocellulose membranes using a mini-blot module with Bolt transfer buffer (ThermoFisher Scientific) for western blotting, or in-gel stained with Pierce Imperial Protein Stain (ThermoFisher Scientific) according to manufacturer's instructions.

Following the protein transfer, the nitrocellulose membranes were blocked in 5% milk powder (Sigma) dissolved in Tris-buffered saline containing 0.1% Tween-20 (TBS-T) for 1h at room temperature.

ZIKV-CON patient B's serum diluted 1:2500 in blocking solution was then allowed to react with the membrane overnight at 4°C. These blots were then washed using 7 changes of TBS-T buffer. A horse-radish peroxidase (HRP)-conjugated Fc-specific anti-human-IgG secondary antibody (A0170, Sigma) was then diluted 1:3000 in blocking solution was then reacted with the membrane for 1h at RT. Detection of the bound IgG autoantibodies was then performed with ECL Prime substrate (GE Healthcare) and the results were developed using radiographic film.

For the subsequent mass spectrometry-based target antigen identification, the bands detected by western blotting were aligned with those in parallel-developed gel stained with the Pierce Imperial Protein Stain (ThermoFisher Scientific). To identify the target protein bound by patient B's IgG autoantibodies gel, the bands identified in the co-culture lysate, along with corresponding positions in the neuron-only and Schwann cell lysates were excised from the chemically stained gel and subjected to in-gel trypsin digestion.

The peptides were then subjected to C18 reverse-phase column chromatography and analysed by data-dependant MS/MS on a ThermoFisher Fusion Lumos mass spectrometer as described above.

**ELISA**

Recombinant human nidogen-1 protein (R&D Systems), laminins (111, 121, 211, 221, 411, 421, 511 and 521) (BioLamina), prosaposin (sulphated glycoprotein-1) (Enzo Life Sciences) and vinculin (VINC, Origene) were prepared according to the manufacturer's instructions and were added at 1 μg/ml (100 μl per well) to Nunc Maxisorp ELISA plates overnight at 4°C (See **Supplementary Table 2** for details). The following day, residual solutions were removed and the wells were blocked with 5% skimmed milk powder (Sigma) in PBS for 1h at room temperature.

Human serum samples were diluted 1:100 in 5% milk were then incubated in the wells for 1h at room temperature. Following five washes with PBS, HRP-conjugated Fc-specific anti-human-IgG secondary antibody (A0170, Sigma) diluted at 1:3000 was added to the wells and incubated for 1h at RT. Following further PBS washing cycles, the bound antibodies were detected by reaction with SigmaFast OPD substrate (Sigma) and terminated after 20 min incubation in the dark at RT with 4M $H_2SO_4$. Optical density measurements were determined at 492 nm and were corrected by subtracting OD values obtained using uncoated wells. The anti-ganglioside GM3 ELISA was performed as previously described (Willison et al., 1999).

For the detection of patients' anti-ZIKV antibodies, Maxisorp (NUNC Immulon 4HB) ELISA plates were coated with the anti-flavivirus monoclonal antibody (4G2) at 2 µg/ml (GTX57154, Genetex) in 0.05 M carbonate-bicarbonate buffer (pH 9.6) overnight at 4°C. ZIKV (PF13 strain) or mock infected C6/36 cells were cultured in Leibovitz L-15 containing 2% fetal bovine serum (FBS), 2 mM L-glutamine, 100 U/ml of penicillin, and 100 µg/ml of streptomycin at 28°C. After 4 days the media supernatants were collected and stored in screw top vials at -80°C (Dejnirattisai et al., 2016).

Duplicate wells were treated with either ZIKV-infected or mock-infected cell culture supernatants for 1 h at 37ºC to capture the Zika virions. After washing the wells with PBS containing 0.1% Tween-20 and blocking them with 3% BSA in PBS, the patients' and healthy control persons' sera diluted at 1:100 in 10% FBS in RPMI were added to the wells and incubated for 1 h at 37ºC. The bound human IgG was then detected with sequential reaction steps with a 1:3000 dilution of an IgG Fc-specific HRP-conjugated secondary antibody (Sigma, A0170) and OPD (Sigma Fast) substrate followed by stopping the reaction with 4M $H_2SO_4$ (see above). The optical density (Absorbance) values were then determined at 492 nm and the resultant anti-ZIKV OD values were determined by subtraction with the OD values obtained from mock-ZIKV treated wells, with the cut-off threshold value of 0.2 for positive reaction determinations.

**Transfected-cell based assays**

HEK293 cells were seeded on 13 mm diameter coverslips coated with PDL (10-25 µg/ml) in medium containing 10% FBS in DMEM with penicillin-streptomycin, L-glutamine and sodium pyruvate. After 24h, these cells were transfected overnight with mammalian expression vectors (0.5-1 µg plasmid cDNA per well) encoding human AHNAK2 (His-tagged), ANXA2 (GFP tagged), CD44S, CNTN1, CNTNAP1 (Caspr1), GFRA1, ITGA6, ITGA7, MAG (mCherry tagged), NEP (MME), NFASC (transcript variants 1 or 2), NKCC1 (SLC12A2) (YFP-tagged), PRX, and TGBR3 (HA-tagged) using JetPEI (Polyplus) (**Supplementary Table 3**). A minimum of 16-24 hours after transfection, human sera were diluted 1:100 in 1% BSA in DMEM containing HEPES and added to the live transfected cells for 1h at RT. Serum-treated cells were then washed in three changes of DMEM/HEPES, and fixed in 4% PFA, before IgG binding was assessed using an Alexa Fluor-488 conjugated goat anti-human IgG (Fc-specific) secondary antibody and fluorescent microscopic analyses. Additionally, human ALCAM, AXL, DPYSL2, GAS6, L1CAM, NCAM1 and NrCAM gene expression plasmids (Origene, Rockville, MD) were transfected at 37°C using Lipofectamine 2000 (Invitrogen) (**Supplementary Table 4**). These cells were subsequently fixed with 4 % PFA, blocked for 1h with 5% NGS and frozen at -80°C until immunocytochemistry experiments were performed. Coverslips containing the transfected HEK293 cells were thawed with 5% NGS. Human sera diluted at 1:100 were added and after incubation the binding of their IgG and IgM autoantibodies were assessed using the method described above for their detection in primary cells. Successful transfection and transgene expression was confirmed by labelling these cells with a commercial antibody, or visualisation of fluorescent tag, and stained coverslips were mounted with Vectashield with DAPI. All plasmid cDNA was sequenced by Sanger Sequencing by a commercial provider (Source Bioscience) and compared to the known open reading frame sequence to prior to transfection assays. The antibody details are described in the **Supplementary Tables 3 & 4**, respectively.

**Statistical analysis**

The results were analysed in Prism v9.1.0 (GraphPad Software). Statistical comparison of the proportions of seropositive patients among the ZIKV groups were performed using contingency analysis with the application of a two-tailed Fisher's exact test for individual group-group comparisons, and Chi square testing for comparisons between all groups. For the fluorescence intensity profiles of the human IgG antibody, human IgM antibody and myelin basic protein (MBP) immunolabelling, confocal images were acquired using identical settings and analysed using the plot profile function on ImageJ. For the analysis of IgM antibody deposition on myelin outpouchings, two images were acquired per serum sample and analysed by an investigator blinded to the sample grouping. IgM antibody fluorescence values for the plot profile were integrated according to the signal overlap with MBP. The outpouching intensity score was background subtracted and normalised to the signal at the adjacent internode. For the analysis of the internodes of solvent or neuraminidase-treated cultures, the images of two internodes per field of view, 6-14 fields of view per treatment from two independent experiments were acquired. Immunofluorescence images were adjusted for brightness and contrast for presentation only. For the comparison of the peptide enrichment in the IP eluates from the ZIKV serum and positive control serum-treated cultures, respectively, was performed on normalised abundance values by t-test using a false discovery rate of 1% and significance threshold of 0.1 (Perseus) (Tyanova et al., 2016). Data were presented as mean ± SEM unless otherwise stated. An alpha-level of $<0.05$ was accepted for statistical significance.

### SUPPLEMENTARY TABLES

**Supplementary Table 1.** Summary of ZIKV patient cohorts and controls.

| Group | Number of samples | Age: median; range [# samples] | %F | ZIKV symptoms to sampling (days) median; range [# samples] | GBS onset to sampling (days) median; range [# samples] |
|---|---|---|---|---|---|
| **ZIKV-GBS (overall)** | **52** | **44; 10-71 [47/52]** | **46.2% [52/52]** | **113; 65-579 [31/52]** | **127; 66-534 [31/52]** |
| • AIDP | 16 | 41; 10-69 [16/16] | 56.3% [16/16] | 102; 67-164 [16/16] | 103; 74-260 [16/16] |
| • AMAN/AMSAN | 10 | 44; 27-70 [10/10] | 50.0% [10/10] | 113; 65-579 [10/10] | 132; 66-300 [10/10] |
| • Unclassified | 26 | 45; 20-71 [21/26] | 38.5% [26/26] | 546; 99-579 [5/26] | 482; 104-534 [5/26] |
| **Controls (overall)** | **225** | **40; 2-88 [221/225]** | **56.9% [225/225]** | **N/A** | **N/A** |
| • ZIKV-OND* | 17 | 26; 8-41 [13/17] | 52.9% [17/17] | 141; 90-313 [12/17] | N/A |
| • ZIKV-CON | 117 | 43; 2-83 [117/117] | 64.1% [117/117] | 105; 1-615 [116/117] | N/A |
| • DENV-CON | 23 | 36; 21-71 [23/23] | 34.8% [23/23] | N/A | N/A |
| • Local healthy* | 9 | 32; 4-41 [9/9] | 55.6% [9/9] | N/A | N/A |
| • ALS | 5 | 68; 35-81 [5/5] | 60.0% [5/5] | N/A | N/A |
| • Dysferlin | 5 | 51; 17-88 [5/5] | 40.0% [5/5] | N/A | N/A |
| • Healthy controls | 49 | 42; 21-80 [49/49] | 55.1% [49/49] | N/A | N/A |

When data was available, the ZIKV infected GBS (ZIKV-GBS) patients were subdivided into those diagnosed as acute inflammatory demyelinating polyneuropathy (AIDP), predominantly axonal (AMAN/AMSAN: acute motor axonal neuropathy or acute motor sensory axonal neuropathy) or ‘unclassified’ where the data were unavailable. Control samples obtained locally included ZIKV patients who presented with other neurological disease (OND*), including myeloradiculopathy (n=1), encephalitis (n=5), meningoencephalitis (n=1), peripheral facial palsy (n=3), transverse myelitis (n=7), uncomplicated ZIKV infections (ZIKV-CON), post-dengue virus infections (DENV-CON) (n=23) and otherwise healthy local controls* (n=9). 38 serum samples from Spanish subjects included patients with the non-immune neuromuscular disorders amyotrophic lateral sclerosis (ALS) (n=5) and dysferlinopathy (Dysferlin) (n=5). 28 healthy Spanish controls were grouped with 21 healthy controls from Oxford. Age and time to sampling data are presented as median, range and the proportion of patients for whom this data was available.

**Supplementary Table 2.** List of candidate antigens evaluated by ELISA.

| Antigen | Coating | Plate | Sera dilution | Blocking solution | Secondary antibody | Secondary antibody dilution |
|---|---|---|---|---|---|---|
| Human Laminins (111, 121, 211, 221, 411, 421, 511, 521) (BioLamina) | 1-5 µg/ml in PBS | Nunc maxisorp | 1:100 | 5% milk in PBS | Anti-human IgG(Fc)-HRP (Sigma, A0170) | 1:3000 |
| Human Nidogen-1 (2570-ND, R&D Systems) | 1-5 µg/ml in PBS | Nunc maxisorp | 1:100 | 5% milk in PBS | Anti-human IgG(Fc)-HRP (Sigma, A0170) | 1:3000 |
| Human Prosaposin / Sulfated glycoprotein-1 (Enzo Life Sciences) | 1 µg/ml in PBS | Nunc maxisorp | 1:100 | 5% milk in PBS | Anti-human IgG(Fc)-HRP (Sigma, A0170) | 1:3000 |
| Ganglioside GM3 from bovine brain (Insight Biotechnology) | 2 µg/ml in methanol | Immulon 2HB | 1:100 | 2% BSA in PBS | Anti-human IgG(Fc)-HRP (Sigma, A0170) | 1:3000 |
| Sulfatides (24323-1 mg-CAY, Cambridge Biosciecnces) | 5µg/ml (in methanol) | Immulon 2HB | 1:100 | 2% BSA in PBS | Anti-human IgG(Fc)-HRP (Sigma, A0170) | 1:3000 |
| Vinculin (Origene) | 1 µg/ml (in carbonate/ bicarbonate buffer) | Nunc maxisorp | 1:100 | 5% milk in PBS + 0.1% Tween-20) | Rabbit anti-human IgG HRP (P0214, Agilent) | 1:3000 |

**Supplementary Table 3.** List of candidate antigens for ZIKV-CON patient B's serum IgG autoantibody evaluated using transfected cell-based assays.

| cDNA clone | Protein of interest | Permea-bilised | Sera dilution | Blocking solution | Positive control antibody | Antibody dilution | Secondary antibodies | Antibody dilution |
|---|---|---|---|---|---|---|---|---|
| pCMV3-AHNAK2-His (HG15220-CH, Sino Biological) Selection: Hygromycin Resistance: Kanamycin | AHNAK2 (Human) | No | 1:200 | 1% BSA in DMEM/ HEPES at 37°C | Rabbit anti-His (2365T, Cell Signalling) | 1:2000 | Goat anti-human IgG (H+L) 488 Goat anti-rabbit IgG Alexa 546 | 1:750, 1:1000 |
| pEGFP-N3-anxA2-GFP (Addgene plasmid #107196) Selection: Neomycin Resistance: Kanamycin (Rescher et al., 2000) | Annexin A2 (Human) | No | 1:100 | 1% BSA in DMEM at 37°C | n/a | n/a | Donkey anti-human IgG (Fab) Cy3 | 1:200 |
| pBABE-puro-CD44S (Addgene plasmid #19127) Selection: Puromycin Resistance: Ampicillin (Godar et al., 2008) | CD44S (Human) | No | 1:100 | 1% BSA in DMEM at 37°C | Sheep anti-CD44 (AF6127, R&D Systems) | 1:1000 | Goat anti-human IgG (H+L) 488 Donkey anti-sheep IgG 546 | 1:750, 1:1000 |
| pRc/CMV-CNTN1 (EX-A1153-M02, Genecopoeia)Selection: Neomycin Resistance: Ampicillin | CNTN1 (Human) | No | 1:200 | 1% BSA in DMEM/ HEPES at 37°C | Goat anti-CNTN1 (AF904, R&D Systems) | 1:2000 | Donkey anti-human IgG (Fab) Cy3 Donkey anti-goat IgG Alexa 488 | 1:200, 1:1000 |
| pReceiver-M02 -CASPR1 (Genecopoeia) and pRc/CMV-CNTN1 EX-A1153-M02, Genecopoeia) | CNTN-1 /Caspr1 (Human) | No | 1:100 | 1% BSA in DMEM/ HEPES | n/a | n/a | Goat anti-human IgG (Fab) 488 | 1:750 |
| pcDNA3.1-GFRA1-C-(K)DYK (OHu12279D, Genscript) Selection: Neomycin Resistance: Ampicillin | GDNF family receptor alpha 1 (Human) | No | 1:100 | 1% BSA in DMEM/ HEPES | Rabbit-anti-FLAG (F7425, Sigma) | 1:500 | Goat anti-human IgG (Fab) 488 Donkey anti-rabbit IgG 546 | 1:750, 1:1000 |
| pTT3-ITGA6-His (Addgene Plasmid #53352) Resistance: Ampicillin (Sun et al., 2015) | ITGA6 (Human) | No | 1:200 | 1% BSA in DMEM/ HEPES at 37°C | Rabbit anti-His (2365T, Cell Signalling) | 1:2000 | Goat anti-human IgG (H+L) 488 Goat anti-rabbit IgG Alexa 546 | 1:750, 1:1000 |
| pCMV3-ITGA7-Myc (HG13425-CM, Stratech) Selection: Hygromycin Resistance: Kanamycin | ITGA7 (Human) | No | 1:100 | 1% BSA in DMEM/ HEPES | Rabbit anti-Myc (A9106, Abcam) | 1:1000 | Goat anti-human IgG (Fab) 488 Donkey anti-rabbit IgG 546 | 1:750, 1:1000 |
| pEZ-M56MAG-mCherry (EX-D0078-M56, GeneCopoeia) Selection: Neomycin Resistance: Ampicillin | MAG (Human) | No | 1:100 | 1% BSA in DMEM at 37°C | n/a | n/a | Goat anti-human IgG (H+L) 488 | 1:750 |
| pBOB-NEP (Addgene plasmid #12338) Resistance: Ampicillin (Marr et al., 2004) | Neprilysin (CD10) (Human) | No | 1:100 | 1% BSA in DMEM at 37°C | Goat anti-NEP (AF1182, R&D Systems) | 1:1000 | Donkey anti-human IgG (Fab) Cy3 Donkey anti-goat IgG Alexa 488 | 1:200, 1:1000 |
| pCMV6-NFASC-MyC-DDK (RC228652, Origene) Selection: Neomycin Resistance: Kanamycin | NFASC 155 (Human) | No | 1:100 | 1% BSA in DMEM/ HEPES | Chicken anti-pan neurofascin (AF3235, R&D Systems) | 1:1000 | Goat anti-human IgG (Fab) 488 Goat anti-chicken IgY Alexa 546 | 1:750, 1:1000 |
| pcDNA3.1 –Nfasc186-Myc Querol lab. Resistance: Ampicillin | NFASC 186 (Human) | No | 1:100 | 1% BSA in DMEM/ HEPES | Chicken anti-pan neurofascin (AF3235, R&D Systems) | 1:1000 | Goat anti-human IgG (Fab) 488 Goat anti-chicken IgY Alexa 546 | 1:750, 1:1000 |
| pcDNA3.1 Flag YFP hNKCC1 (Addgene plasmid #49085) Selection: Neomycin Resistance: Ampicillin (Somasekharan et al., 2013) | NKCC1 (Human) | No | 1:200 | 1% BSA in DMEM/ HEPES at 37°C | n/a | n/a | Donkey anti-human IgG (Fab) Cy3 | 1:200 |

| | | | | | | | | |
|---|---|---|---|---|---|---|---|---|
| pcDNA3.1-PRX_tv2-C-(K)DYK (OHu25883, Genscript) Selection: Neomycin Resistance: Ampicillin | L-Periaxin (PRX) (Human) | No/Yes | 1:100 | 1% BSA in DMEM/ HEPES | Sheep anti-PRX (gift from P. Brophy) | 1:2000 | Goat anti-human IgG (Fab) 488 Donkey anti-sheep IgG 546 | 1:750, 1:1000 |
| pcDNA3.1-PRX_tv1-C-(K)DYK (OHu25782, Genscript) Selection: Neomycin Resistance: Ampicillin | S-Periaxin (PRX) (Human) | Yes | 1:100 | 1% BSA in DMEM/ HEPES | Rabbit anti-FLAG (F7425, Sigma) | 1:500 | Goat anti-human IgG (Fab) 488 Donkey anti-rabbit IgG 546 | 1:750, 1:1000 |
| pBABE-neo-TGFBR3-HA (Addgene plasmid #83095) Selection: Puromycin Resistance: Ampicillin (Bajikar et al., 2017) | TGFBR3 (Human) | No | 1:100 | 1% BSA in DMEM at 37°C | Rabbit anti-HA (C29F4) (#3724, CST) | 1:1000 | Goat anti-human IgG (H+L) 488 Goat anti-rabbit IgG Alexa 546 | 1:750, 1:1000 |

### Abbreviations

AHNAK2, AHNAK nucleoprotein 2 (617 kDa)
ANXA2 Annexin-A2 (40 kDa)
BSA, bovine serum albumin
DDK/DYK, FLAG-tag (DYKDDDDK sequence motif)
DMEM, Dulbecco's Modified Eagles Medium
GDNF, Glial-derived neurotrophic factor
GFRA1 GDNF family receptor alpha 1
HEPES, 2-[4-(2-hydroxyethyl)piperazin-1-yl]ethanesulfonic acid
ITGA6 Integrin alpha 6 (127 kDa)
ITGA7 Integrin alpha 7 (130 kDa)
MAG, Myelin-associated glycoprotein (70 kDa)
NEP, Neprilysin, membrane metallo-endopeptidase (90 kDa)
NFASC, Neurofascin (155 kDa and 186 kDa)
NGS, normal goat serum.
NKCC1 (SLC12A2), Sodium potassium chloride co-transporter 1 (130 kDa)
PRX L-Periaxin (155 kDa)
PRX S-Periaxin (16 kDa)
TGFBR3, Transforming growth factor receptor beta 3 (94 kDa)
YFP, Yellow fluorescent protein

### Secondary antibody details:

Goat anti-human IgG (Fab) Alexa 488 (H10120, Life Technologies)
Goat anti-human IgG (H+L) Alexa 488 (A11013, Life Technologies)
Goat anti-human IgG Alexa 568 (A21090, Life Technologies)
Goat anti-chicken IgY Alexa 546 (A11040, Life Technologies)
Goat anti-mouse IgG Alexa 546 (A11003, Life Technologies)
Goat anti-rabbit IgG Alexa 546 (A11010, Life Technologies)
Donkey anti-human IgG (Fab) Cy3 (709-166-098, Jackson)
Donkey anti-sheep IgG Alexa 546 (A21098, Life technologies)
Donkey anti-goat IgG Alexa 488 (A11055, Life Technologies)
Donkey anti-rabbit IgG Alexa 546 (A10040, Life Technologies)

**Supplementary Table 4.** List of candidate antigens for patient C (IgG) and patient D (IgG/IgM) autoantibodies evaluated by transfected cell-based assays**.**

| cDNA clone | Protein of interest | Permea-bilised | Sera dilution | Blocking solution | Positive control antibody | Antibody dilution | Secondary antibodies | Antibody dilution |
|---|---|---|---|---|---|---|---|---|
| pCMV6-ALCAM-Myc-DDK (RC219251, Origene) | ALCAM (Human) | No | 1:100 | 5% NGS in PBS | Mouse-anti-Myc (CBL430 Millipore) | 1:200 | Goat anti-mouse IgG 488, Goat anti-human IgG 594 | 1:1000, 1:1000 |
| pCMV6-AXL-MyC-DDK (RC206431, Origene) Selection: Neomycin Resistance: Kanamycin | AXL (Human) | Yes | 1:100 | 5% NGS in PBS | Mouse anti-Myc (CBL430 Millipore) | 1:200 | Goat anti-mouse IgG 488, Goat anti-human IgG 594 | 1:1000, 1:1000 |
| pReceiverM02-DPYSL2 (EX-M0208-M02-10, Genecopoeia) | DPYSL2 (Human) | Yes | 1:100 | 5% NGS in PBS | Rabbit anti-DPYSL2 (ab129082 Abcam) | 1:100 | Goat anti-rat IgG 488, Goat anti-human IgG 594 | 1:1000, 1:1000 |
| pCMV6-GAS6-MyC-DDK (RC207916, Origene) Selection: Neomycin Resistance: Kanamycin | GAS6 (Human) | Yes | 1:100 | 5% NGS in PBS | Mouse anti-Myc (CBL430 Millipore) | 1:200 | Goat anti-mouse IgG 488, Goat anti-human IgG 594 | 1:1000, 1:1000 |
| pCMV6-L1CAM-Myc-DDK (RC211601, Origene) | L1CAM (Human) | Yes | 1:100 | 5% NGS in PBS | Mouse anti-Myc (CBL430 Millipore) | 1:200 | Goat anti-mouse IgG 488, Goat anti-human IgG 594 | 1:1000, 1:1000 |
| pCMV6-NCAM1-C-His (EX-X0019-M77 Genecopoeia) | NCAM1 (Human) | No | 1:100 | 5% NGS in PBS | Mouse anti-NCAM (347740 BD Biosciences ) | 1:1000 | Goat anti-mouse IgG 488, Goat anti-human IgG 594 | 1:1000, 1:1000 |
| pCMV6-NrCAM (EX-H0649-M02, Genecopoeia) Selection: Neomycin Resistance: Ampicillin | NrCAM (Human) | Yes | 1:100 | 5% NGS in PBS | Rat anti-HA (supplier) | 1:20000 | Goat anti-rat IgG 488, Goat anti-human IgG 594 | 1:1000, 1:1000 |

### Abbreviations

ALCAM (CD166), Activated leukocyte cell adhesion molecule (65 kDa)
AXL, Tyrosine-protein kinase receptor UFO (98 kDa)
BSA, bovine serum albumin.
DMEM, Dulbecco's Modified Eagles Medium
DPYSL2, Dihydropyrimidinase-related protein 2 (62 kDa)
GAS6, Growth arrest-specific protein 6 (75kDa)
L1CAM, Neural cell adhesion molecule L1 (140 kDa)
NCAM1 Neural cell adhesion molecule 1 (94 kDa)
NGS, normal goat serum.

NrCAM, Neuronal cell adhesion molecule (143 kDa)

**Secondary antibody details:**

Goat anti-human IgG Alexa 594 (Ab98621 Abcam)
Goat anti-mouse IgG Alexa 488 (A11001, Life Technologies)
Goat anti-rat IgG Alexa 488 (A11008, Invitrogen)

**Supplementary Table 5.** List of candidate antigens identified by immunoprecipitation and mass spectrometry, or literature review.

| Target antigen | Abbreviation | Function | Supplementary data file |
|---|---|---|---|
| AHNAK2 | AHNAK2 | A scaffolding or nucleoprotein previously designated C14orf78 (also known as desmoyokin) is reported to interact with the myelin protein periaxin (Han and Kursula, 2014), and is closely related to AHNAK1, which is thought to enhance attachment of the abaxonal membrane to the lamina substrate in myelinating Schwann cells (Salim et al., 2009). | Patient B IgG - myelinating co-cultures (Data 2) |
| Activated leukocyte cell adhesion molecule | ALCAM (CD166) | Cell adhesion molecule found on blood–brain barrier endothelial cells (BBB-ECs) that was previously shown to be involved in leukocyte transmigration across the endothelium, and has an important function in maintaining BBB integrity. (Lecuyer et al., 2017). | Patient C IgG - DRG neurons (Data 3) Patient D IgM - DRG neurons (Data 5) |
| Annexin A2 | ANXA2 | One of a family of phospholipid binding proteins that was originally identified as an endothelial cell receptor for plasminogen (Hajjar et al., 1996). ANXA2 is also highly expressed in peripheral nerve myelin (Hayashi et al., 2007). Schwann cells respond to nerve injury by cytoskeletal remodelling and local translation of ANXA2 (Negro et al., 2018), which forms a complex at the plasma membrane with calcium-binding protein S100 (Liu et al., 2015). High levels of anti-ANXA2 antibodies have been positively associated with disease severity in COVID-19 patients (Zuniga et al., 2021). | Patient B IgG - myelinating co-cultures (Data 2) |
| Tyrosine-protein kinase receptor UFO | AXL | Cell surface receptor by which ZIKV entries at human neural progenitor cells (hNPC), leading to a stimulation of AXL-mediated signalling pathways and suppression of the innate immune response. This leads to viral infection in neurons and their associated cells (Faizan et al., 2016; Meertens et al., 2017). | Literature review |
| P-glycoprotein 1 | CD44 | A cell-surface glycoprotein involved in cell-cell adhesion between Schwann cells and neurons (Sherman et al., 2000). | Patient B IgG - myelinating co-cultures (Data 2) |
| Contactin 1 | CNTN1 | Protein involved in the formation of paranodal axo-glial junctions in myelinated peripheral nerves and in the signaling between axons and myelinating glial cells via its association with CASPR1 (Rios et al., 2000). | CNTN1+ IgG – iPSC derived neuronal cultures (Data 1) Patient D IgM - DRG neurons (Data 5) |
| Dihydropyrimidinase-related protein 2 | DPYSL2 | Plays a role in neuronal development and polarity, as well as in axon growth and guidance, neuronal growth cone collapse and cell migration (Inagaki et al., 2001). | Patient D IgG - DRG neurons (Data 4) Patient D IgM - DRG neurons (Data 5) |
| Growth arrest-specific protein 6 | GAS6 | Ligand of AXL. AXL-mediated ZIKV entry requires the AXL ligand Gas6 to serve as a bridge linking ZIKV particles to glial cells (Meertens et al., 2017) | Literature review |
| GDNF family receptor alpha-1 | GFRA1 | Receptor component for glial cell line-derived neurotrophic factor. Found within the oute rmembrane of the myelin sheath of peripheral nerves (Hase et al., 2005). | Patient B IgG - myelinating co-culture lysate (Western blot) (Data 6) |

| | | | |
|---|---|---|---|
| Integrin alpha 6 | ITGA6 | The integrin alpha 6 subunit associates with beta 4, which together play a role in Schwann cell myelination via interactions with the basal lamina (Einheber et al., 1993). | Patient B IgG - myelinating co-cultures (Data 2) |
| Integrin alpha 7 | ITGA7 | Expressed in postnatal Schwann cells, which it is thought to bind laminins within the endoneurium and may play a role in nerve regeneration (Previtali et al., 2003). | Patient B IgG - myelinating co-culture lysate (Western blot) (Data 6) |
| Neural cell adhesion molecule L1 | L1CAM | Neural cell adhesion molecule expressed at the axonal surface. In the PNS, it has been shown that L1CAM is involved in the myelination process, as antibodies against this protein inhibit Schwann cells myelination of dorsal root ganglia neurons (Coman et al., 2005). | Patient C IgG - DRG neurons (Data 3) Patient D IgM - DRG neurons (Data 5) |
| Myelin associated glycoprotein | MAG | A transmembrane glycoprotein in myelinating Schwann cells (Quarles, 2007) that also inhibits neurite outgrowth after injury (Mukhopadhyay et al., 1994) and Schwann cell migration (Chaudhry et al., 2017) | Literature review |
| Neural cell adhesion molecule 1 | NCAM1 | Cell adhesion molecule involved in neuron-neuron adhesion, neurite fasciculation, outgrowth of neurites, etc. Recently it has been demonstrated to act as a receptor for ZIKV (Srivastava et al., 2020). | Patient D IgM - DRG neurons (Data 5) |
| Neural cell adhesion molecule 2 | NCAM2 | NCAM2 plays a key role in neurodevelopment. Expression is down-regulated following Zika virus infection (Barbeito-Andres et al., 2020). | Patient A IgG - iPSC derived neuron cultures (Data 1) |
| Neprilysin | NEP (MME) | Also known as membrane metallo-endopeptidase (MME) and cluster of differentiation 10 (CD10). Mutations of this gene have been associated with Charcot-Marie-Tooth Type 2 (CMT2) and axonal polyneuropathies (Auer-Grumbach et al., 2016). | Patient B IgG - myelinating co-cultures (Data 2) |
| Neurofascin | NFASC | Cell adhesion molecule with neuronal (NF186) and glial (NF155) isoforms sharing a common extracellular immunoglobulin domain. Critical for glial attachment to axons and clustering of sodium channels at the node of Ranvier (Lustig et al., 2001). | Patient A IgG - iPSC derived neuron cultures (Data 1) |
| Neuronal cell adhesion molecule | NrCAM | Neuronal cell adhesion molecule that plays a role in the formation and maintenance of the nodes of Ranvier on myelinated axons. This protein shares 6 peptides with ZIKV polyprotein (Lucchese and Kanduc, 2016). | Literature review |
| Solute carrier Na-K-2Cl cotransporter 1 | NKCC1 (SLC12A2) | Plasma membrane co-transporter of Na+, K+, and Cl− ions and water, typically from the extracellular space into cells. Regulates ion homeostasis in myelinating Schwann cells following neuronal activity (Marshall-Phelps et al., 2020). | Patient B IgG - myelinating co-cultures (Data 2) |
| Periaxin | PRX | PDZ domain protein involved in myelin sheath stabilization. The long form (L-PRX) is located at the plasma membrane of myelinating Schwann cells, whereas the short form (S-PRX) is found diffusely throughout the cytoplasm (Dytrych et al., 1998). The distribution of L-PRX is highly dynamic during development and after nerve injury (Scherer et al., 1995). | Patient B IgG - myelinating co-culture lysate (Western blot) (Data 6) |
| Prosaposin | PSAP | A membrane glycoprotein precursor for Saposins (also known as sulfated glycoprotein 1). described as a lysosomal or secreted Originally described as a binder of a-series gangliosides (Hiraiwa et al., 1992). May have a pro-myelinating regenerative role in secreted form after peripheral nerve injury (Hiraiwa et al., 1999) and increases | Literature review |

| | | | |
|---|---|---|---|
| | | sulfatide production by Schwann cells via a G-protein/MAPK dependent mechanism (Campana et al., 1998). Prosaposin has also been detected on the plasma membrane of neuroblastoma cells (Fu et al., 1994) where it forms a complex with GM3 ganglioside (Misasi et al., 1998). | |
| Transforming growth factor receptor type 3 | TGFBR3 | Also known as Betaglycan, binds to the TGF-B superfamily of ligands. Expression in Schwann cells is regulated in response to forskolin treatment (Schmid et al., 2014). | Patient B IgG - myelinating co-cultures (Data 2) |
| Vinculin | VINC | Actin filament binding protein involved in cell-matrix adhesion and cell-cell adhesion (Carisey and Ballestrem, 2011). It has been described as a target antigen in sera from 2 chronic inflammatory demyelinating polyneuropathy (CIDP) patients (Beppu et al., 2015) | Patient C IgG - DRG neurons (Data 3) Patient D IgG - DRG neurons (Data 4) |
